# Hair salons as a promising space to provide HIV and sexual and reproductive health services for young women in Lesotho: A citizen scientist mixed-methods study

**DOI:** 10.1101/2024.12.16.24318906

**Authors:** Malena Chiaborelli, Mamaswatsi Kopeka, Pontšo Sekhesa, Madeleine Sehrt, Tsepang Mohloanyane, Tala Ballouz, Dominik Menges, Jennifer A. Brown, Jennifer M. Belus, Felix Gerber, Fabian Raeber, Andréa Williams, David Jackson-Perry, Meri Hyöky, Donaldson F. Conserve, Karen Hampanda, Alain Amstutz, the Hair SALON Citizen Scientist Working Group

**Author notes:** shared first authors (Malena Chiaborelli, Mamaswatsi Kopeka). shared last authors (Karen Hampanda, Alain Amstutz). Email addresses: MK PS MS TM TB DM JAB JMB FG FR AW DJP MH DFC KH AA.

## Abstract

**Introduction:** Adolescent girls and young women in southern Africa are disproportionately affected by HIV and sexual and reproductive health (SRH) challenges. There is a need for more accessible and de-medicalized community spaces to offer HIV/SRH services for this key population. We aimed to assess the acceptability and feasibility of offering HIV/SRH services at hair salons in Lesotho.

**Methods:** We used an innovative citizen scientist mixed-methods approach, whereby hair stylists were recruited through social media, completed questionnaires, and recruited women clients aged 15-35 years as respondents. A stepwise verification process including GPS, pictures, and a local mobile payment system ensured data quality. Subsequently, we conducted individual in-depth interviews among 14 stylists and clients, following the rapid thematic analysis framework, supported by natural language processing. Clients and stylists were involved at the design, implementation, and results interpretation stage.

**Results:** We recruited 157 hair stylists (median age 29; [interquartile range 25-33]; across all ten districts of Lesotho) and 308 women clients (median age 26 [22-30]). Among stylists, 93.6% were comfortable offering oral HIV self-testing (HIVST), 92.4% pre-exposure prophylaxis (PrEP), and 91.7% post-exposure prophylaxis (PEP). Among clients, 93.5%, 88.3%, and 86.4% felt comfortable receiving the above-mentioned services at a hair salon, respectively. Immediate demand was 30.8% for HIVST, 22.1% for PrEP, and 14.9% for PEP. Acceptability and demand were higher for family planning methods and menstrual health products. 90.4% of stylists thought that offering HIV/SRH services would positively impact their business. The majority of clients visit their salon once or twice a month. Salons were more accessible than the nearest health facility in terms of cost and time, but only 21.0% have an additional confidential space. Qualitative analysis confirmed high acceptability of hair salons as an accessible, less judgemental space than clinics, but raised concerns regarding confidentiality and stylists’ roles.

**Conclusions:** This study suggests that offering HIV/SRH services in hair salons in Lesotho is largely acceptable and feasible with some addressable barriers. A pilot intervention, based on recommendations from this study, is warranted to translate these findings into practice.

## Introduction

Young women in southern and eastern Africa are at the epicenter of the HIV pandemic due to a number of biological, social, and structural factors[1]. In 2023, young women and girls in these regions accounted for 63% of all new global HIV acquisitions with more than 4000 acquiring HIV every week[2]. As in other settings with a high HIV prevalence in southern Africa, HIV and pregnancy-related complications are the leading causes of death among young women in Lesotho[3]. Nearly a quarter of the adult population in Lesotho lives with HIV and two-thirds of new HIV acquisitions are among young women[4, 5]. Similarly, high rates of other sexually transmitted infections and unintended pregnancies have been reported in this key population[6, 7].

In 2016, the Lesotho Ministry of Health introduced oral pre-exposure prophylaxis (PrEP) as a prevention strategy into the national HIV program, and as of 2020, promotes broad eligibility criteria with young women being a key population[8, 9]. Despite significant investment in the HIV programme across the region, uptake and continuation of PrEP among young women has been challenging[10]. Providing HIV services to young women is most promising if integrated with other sexual and reproductive health (SRH) services as recommended by the World Health Organization (WHO)[11]. The Christian Health Association of Lesotho, serves an estimated 40% of the population, but provides only limited SRH services[12]. Furthermore, access to traditional health facilities is a major challenge with 70% of Basotho living in rural areas[13]. Innovative community-based integrated HIV/SRH care models are needed to address access barriers and expand HIV/SRH services.

Hair salons may offer a novel and accessible community space to engage young women and address some barriers related to SRH. Women, across all levels of society, spend substantial time in hair salons without male partners, during which they focus on health- and beauty-related issues, receive community news, and socialize. Stylists are often trusted members of the community. While there is promising evidence from the United States that hair salons can improve access to certain health services[14-19], evidence from Africa is limited to four small-scale published surveys and qualitative analyses from Durban, South Africa [20-23].

Citizen science, i.e., involving citizens at various research stages, originates from environmental health research[24] with the aim to foster a deeper public engagement with science, tailor the research agenda to the needs of the public, and democratize science. Citizen science is becoming an increasingly important approach across various research fields[25-27], including HIV implementation science[28].

Using a citizen science approach, we conducted a mixed-methods study among hair salon stylists and women clients to assess the acceptability and feasibility of offering various HIV/SRH services at hair salons across Lesotho.

## Methods

### Study design and setting

This was an explanatory sequential mixed-methods study[29] with an emphasis on the quantitative component, which was used to inform the development of the qualitative component. The quantitative component consisted of an online nationwide cross-sectional survey administered between March 2024 and July 2024. The qualitative part entailed in-depth individual interviews, conducted between May 2024 to July 2024, following a rapid thematic analysis approach[30]. The study protocol was registered [31]. We report the study according to the Mixed Method Reporting in Rehabilitation & Health Services (MMR-RHS) checklist[29] (Text S1) and the community and citizen scientist involvement according to the Guidance for Reporting Involvement of Patients and the Public (GRIPP2) checklist[32] (Text S2).

### Citizen-scientist survey

For the survey, we first recruited citizen scientists, i.e., hair stylists across the entire country, using social media and a dedicated registration webpage (https://hairsalonproject.com/). Inclusion criteria for stylists were being aged 18 years or older, working at a women’s hair salon in Lesotho (not house call service only), owning an Android smartphone with a functional camera and GPS, using WhatsApp, and having an M-PESA account (a widely used mobile money service in Lesotho). Once successfully registered (i.e., eligibility checked, existence of the hair salon verified, and electronic consent given), the citizen scientists received a link to their questionnaire via WhatsApp and were subsequently asked to recruit their clients. Inclusion criteria for clients were being 15 to 35 years old, self-identifying as women, being clients of the participating stylists, owning an Android smartphone with a camera and GPS, using WhatsApp, and having an M-PESA mobile account. Stylists provided their recruited clients with a link to the same registration webpage. After successful registration on the webpage and providing electronic consent, the clients received their questionnaire via WhatsApp. At each step, the data was centrally verified by the study coordinator in real-time, using various verification criteria (details in Figure S1).

To quantitatively assess the acceptability of offering/receiving HIV/SRH services among stylists/clients, we used three different measures. First, we asked stylists and clients to rate their level of comfortability of offering/receiving thirteen different HIV/SRH services (Text S3) on a 5-level likert-scale (strongly disagree to strongly agree). Second, we asked the clients which of the thirteen services they would be interested in receiving if provided at the hair salon within the coming 6 months. Third, we asked the stylists if they believed providing any of these services would have a positive/negative impact on their business (questions on a 5-level likert-scale). Feasibility considerations in the stylists’ questionnaire encompassed the salon’s infrastructure, opening hours, staff and client volume, as well as reimbursement expectations. The clients’ feasibility questionnaire section entailed the frequency, duration and resource use of a typical salon visit versus a typical health facility visit. Among all participants, we collected socio-demographic data, awareness of HIV self-testing (HIVST) and PrEP, PrEP myths and perceptions (adapted from[33]), age of menarche, menstrual material use[34], impact of menstruation on work/school attendance, current family planning methods use and supply location, and clients’ own perceived HIV risk (one-question risk index from prior research[35]). We collaboratively developed the questionnaires in English and Sesotho and piloted them during a citizen scientist workshop (Text S2). The questionnaires are available in the supplement (Text S4 and S5). The questionnaires were deployed through the web interface of KoboCollect[36, 37], transferring the data via a secured channel onto a password-protected server. Using an application programming interface via KoboconnectR[38] and a project dashboard based on shinyR[39], the study coordinator monitored and verified the data real-time.

### Individual in-depth interviews

For individual in-depth interviews, we recruited survey participants (stylists and clients) using purposive sampling to achieve a broad spectrum of perceptions while reaching thematic saturation. Based on data from the survey, we specifically invited stylists and clients with opposing views (e.g. not comfortable offering/receiving certain HIV/SRH services vs very comfortable offering/receiving certain HIV/SRH services) or who were critical about the project. All interested individuals were sent a digital copy of the consent form in their preferred language prior to the set interview date. The interview guide was developed collaboratively with the Citizen Scientist Working Group (Text S2). The aim was to explore five domains: i) perception of offering/receiving each of the 13 HIV/SRH services at hair salons (Text S6), ii) facilitators of offering/receiving any HIV/SRH service at hair salons, iii) barriers to offering/receiving any HIV/SRH service at hair salons, iv) general HIV/SRH knowledge, access, uptake and stigma, and v) the involvement in research as citizen scientists (among stylists only). The interviews were conducted in-person, at a confidential and comfortable space according to participants’ preferences. Participants were compensated for their time and travel costs between 50 to 100 LSL (2.9 to 5.8 USD), depending on their travel distance. All interviews were done in English or Sesotho, facilitated by a trained qualitative researcher, herself a young woman from Lesotho, fluent in both English and Sesotho, audio-recorded, and subsequently translated and transcribed into English. We collaboratively developed a rapid thematic analysis matrix[30] with pre-specified themes across the five above-mentioned overarching domains.

### Quantitative and qualitative sample sizes, data analyses, and triangulation

Sample size calculations followed corresponding guidance for cross-sectional cluster surveys[40] and were based on the only available quantitative evidence[21] indicating that 77% of clients were comfortable receiving PrEP at a hair salon in South Africa. Based on this, we calculated that a sample size of at least 300 clients across 100 stylists (with an intra-cluster correlation coefficient [ICC] of 0.05, a commonly recommended ICC for PrEP uptake[41, 42]) would be required to estimate the proportion of clients being comfortable receiving PrEP at hair salons in Lesotho with a 5% margin of error for a 95% confidence interval (details available on <u>GitHub</u>). Following guidance for sample size in qualitative research[43], we planned 15 interviews or until reaching thematic saturation during the rapid thematic analysis.

The survey data was analyzed using appropriate descriptive statistics (frequency and interquartile ranges, mean/median and standard deviation). We stratified all client data analyses by age 15-24 years and above, and collapsed the 5-level likert scales throughout into three categories (i.e., combining “strongly agree” with “agree”, “strongly disagree” with “disagree”, and “neither agree nor disagree” as “neutral”). Data management and analysis was conducted using R version 4.3.3 [44].

The qualitative data analysis, following rapid thematic analysis approach guidance[30], was conducted in English and supported by Avidnote, an academic artificial intelligence tool for natural language processing with secured and regulatory-compliant data storage. We developed prompts for Avidnote to analyze each uploaded transcript to a) summarize the participants’ perception on each of the five domains (see above; last domain only among stylists) and b) pull illustrative quotes for each domain as well as for each of the individual 13 HIV/SRH services assessed in the survey. The results were fed into the pre-specified analysis matrix with deductive codes and additional codes were added inductively. This process was piloted with sample interviews and the prompts were refined after the first transcript that was in parallel independently analyzed manually by MK and MS to establish intercoder reliability.

We triangulated the qualitative data from domains (i) to (iii) with the quantitative data to achieve our aim of assessing acceptability and feasibility of offering/receiving HIV/SRH services at hair salons as comprehensively as possible. The data from domains (iv) and (v) will be reported elsewhere.

### Ethics statement

The study protocol was approved by the National Health Research Ethics Committee in Lesotho (ID 61-2024; approval for the survey on March 1st, 2024; approval for the amendment defining in more detail the qualitative part on April 8th, 2024), including the request for a waiver of parental consent for survey participants aged 15-18 years of age. All participants, for both quantitative and qualitative components, provided prior written informed consent.

## Results

### Study participant characteristics

Between March and July, 2024, 157 stylists and 308 clients successfully registered, completed the questionnaire, and were eligible (flowchart in Figure S2). We recruited hair salons/stylists from all ten districts of the country, though most salons were located in urban and suburban areas with the majority (68.2%) concentrated in the capital Maseru (Figure S3). Stylists recruited a median of 3 clients (range: 1–5).

Most of the stylists identified as women (92.4%) with a median age of 29 years (interquartile range [IQR] 25, 33) (Table 1). All clients identified as women, had a median age of 26 years (IQR 22, 30), and 128 of 308 (41.6%) were 15-24 years old (subsequently termed “younger clients”). Secondary or higher education was completed by 82.2% of stylists, 91.4% of younger clients, and 86.6% of older clients. 6.2% of the younger clients were married and 42.5% of the older clients. Economically, only 10.9% of younger clients and 23.3% of older clients reported their financial situation being “comfortable” or “very comfortable”. 2.5%, 0.8%, and 1.1% reported living with HIV among the stylists, younger clients and older clients, respectively. Among stylists not living with HIV and reporting a self-perceived HIV acquisition risk, 25.6% indicated medium-high risk. This was the case for 19.6% of the younger and 29.9% of the older clients. The baseline characteristics of the 14 interviewees were representative of the survey participants and included survey participants with differing views about offering/receiving HIV/SRH services at the hair salon (<u>Table S1</u>).

**Table 1.**
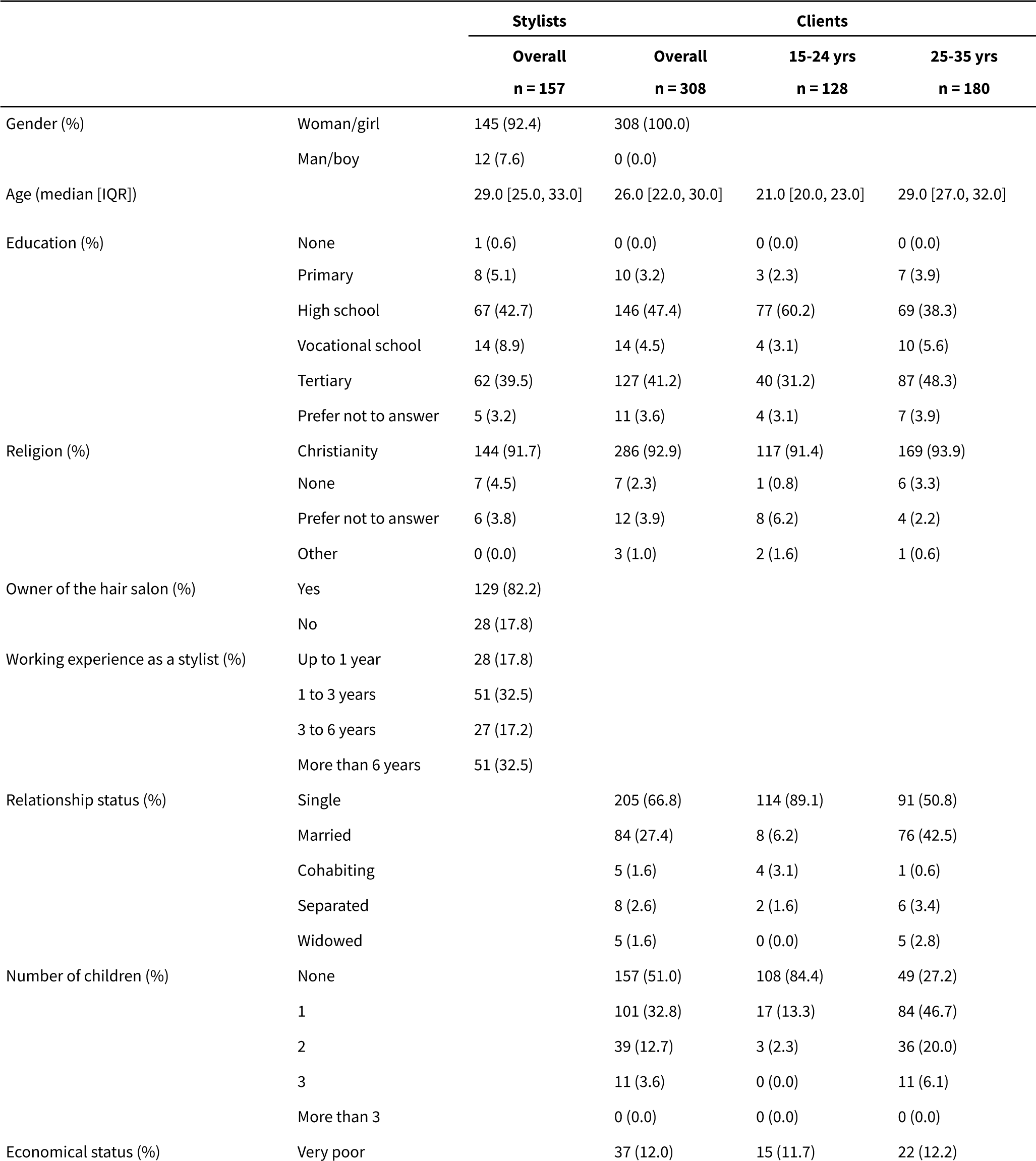
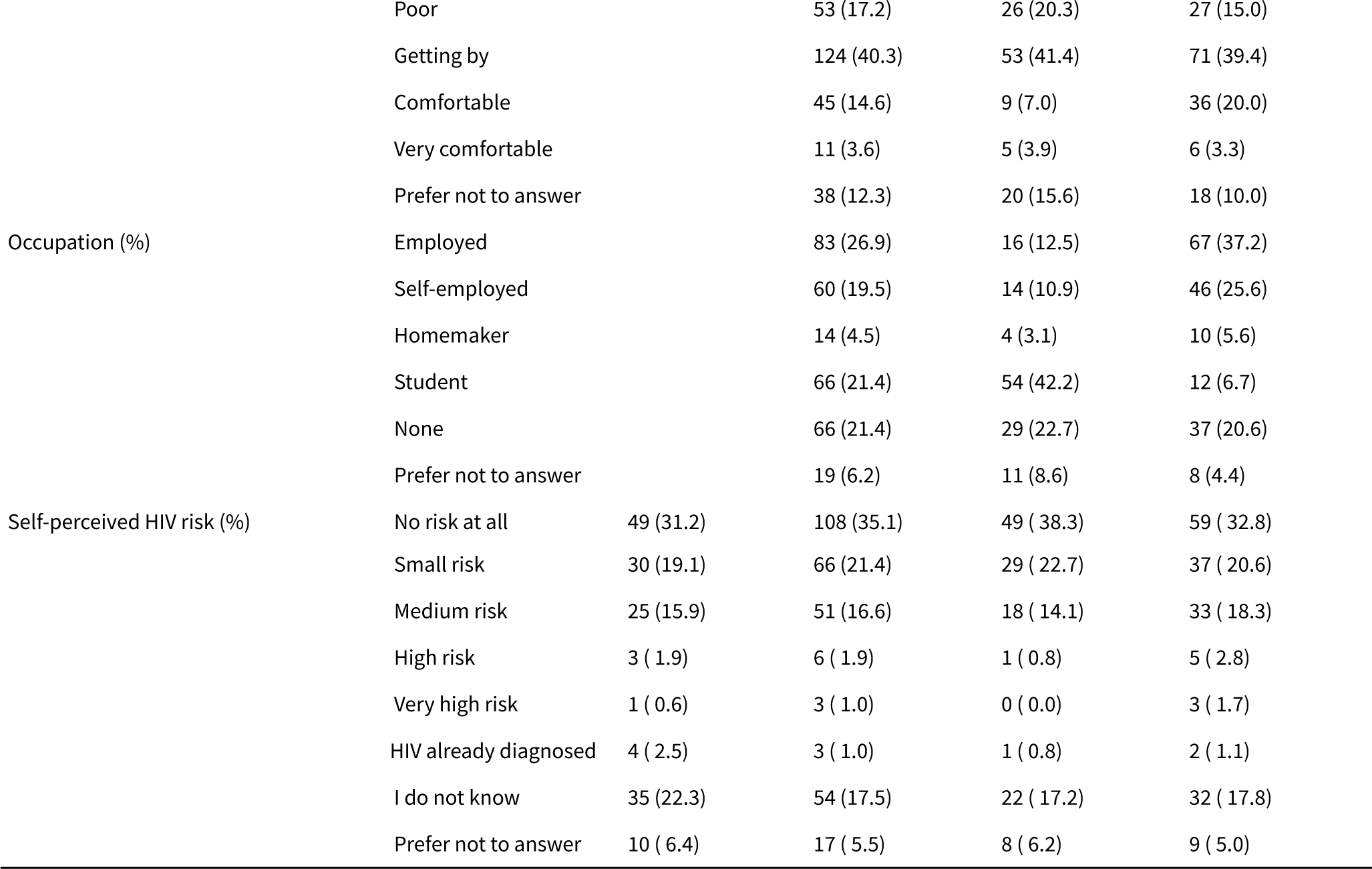
Stylists and clients’ baseline characteristics.

### PrEP, family planning, and menstrual health

Among stylists, 97.5% had heard of HIVST and 91.7% of PrEP; among clients, this was the case for 96.4% and 82.8%, respectively (Table S2). PrEP myths varied with 41.0% of stylists and 42.7% of clients supporting the statement that “PrEP will cause people to have more risky sex”. 63.1% of stylists reported currently using a contraceptive method (mainly male/external condom (32.3%) and the birth control pill (31.3%), most often obtained at a health facility), while 54.9% of clients reported currently using a contraceptive method (mainly male/external condom (45.0%), most often obtained at a health facility). Across all participants who experienced menstruation, disposable sanitary pads were the most commonly used menstrual health product (>90%). 29.5% of stylists and 24.7% of clients mentioned having missed at least one day of school or work because of menstruation in the past year, mainly due to period pain.

### Acceptability of providing HIV/SRH services at hair salons

Among stylists, 86.6% (95% CI: 0.81, 0.91) agreed or strongly agreed to being comfortable offering HIV counseling (Figure 1, Tables S3). Comfortability levels for HIVST, PrEP and PEP were 93.6% (95% CI: 0.90, 0.97), 92.4% (95% CI: 0.88, 0.97), and 91.7% (95% CI: 0.87, 0.96), respectively. For family planning services (counseling, external/male condoms, internal/female condoms, contraceptive pill, emergency contraceptive pill), providing GBV information, and menstrual health services (counseling, sanitary pad provision) the proportion of stylists being comfortable offering these services was generally higher than for HIV-related services, ranging up to 96.2% for menstrual products.

**Figure 1.**
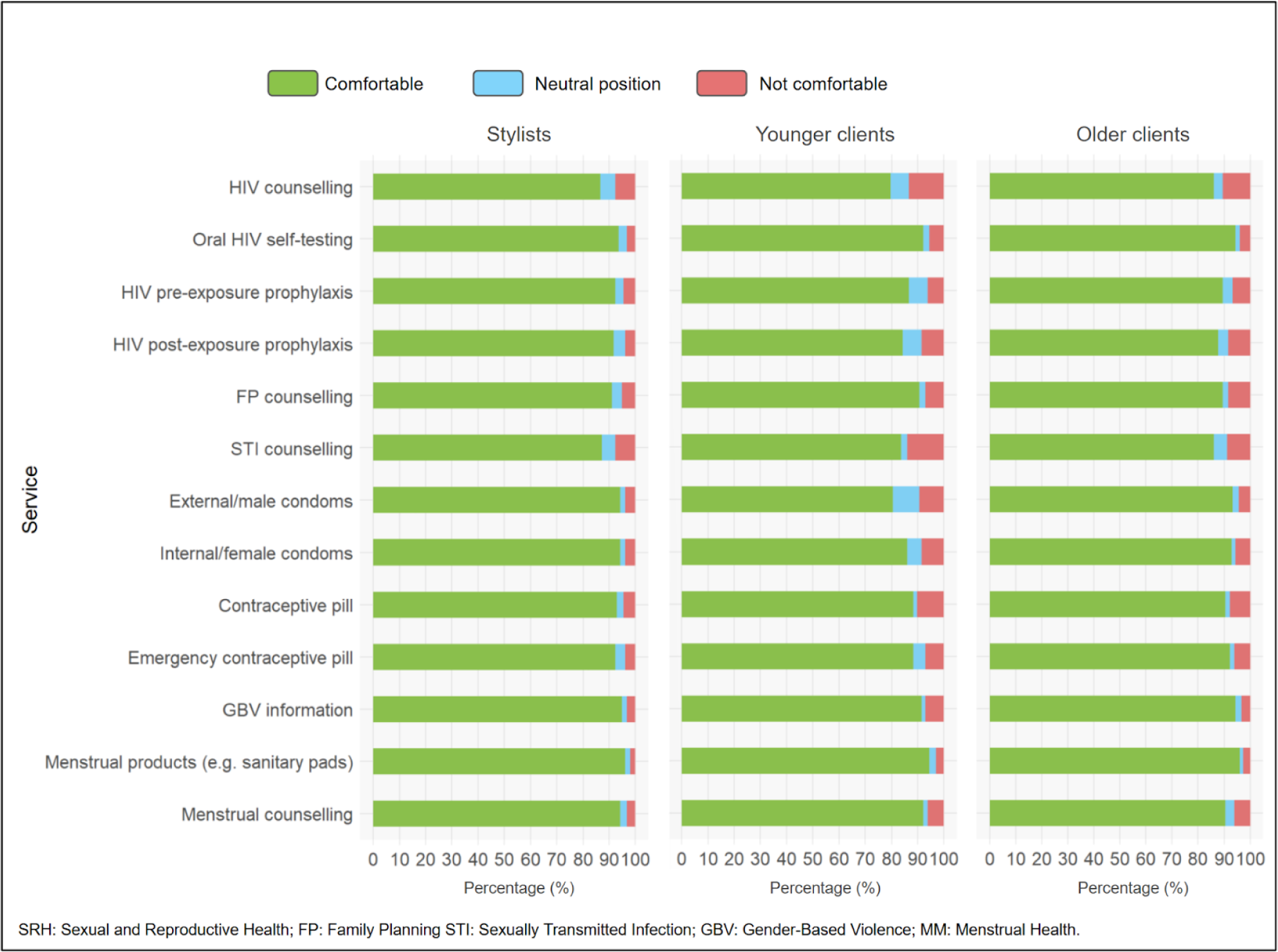
Comfortability of stylists to offer, younger clients and older clients to receive HIV/SRH services at hair salons in Lesotho.

Among younger and older clients, comfortability levels showed a similar pattern, but were overall slightly lower than among stylists, with 83.4% (95% CI: 0.79, 0.88) for HIV counseling, 93.5% (95% CI: 0.91, 0.96) for HIVST, 88.3% (95% CI: 0.85, 0.92) for PrEP and 86.4% (95% CI: 0.83, 0.90) for PEP (Figure 1, Table S4).

The interviews provided rich insights regarding barriers and facilitators of offering/receiving HIV/SRH services at hair salons. Figure 2 summarizes the codes and table S5 additionally the themes. Besides accessibility and convenience arguments outlined in the next section, stylists and clients mentioned themes like “safe space” and “non-judgemental” as facilitators of providing these services at hair salons:

**Figure 2.**
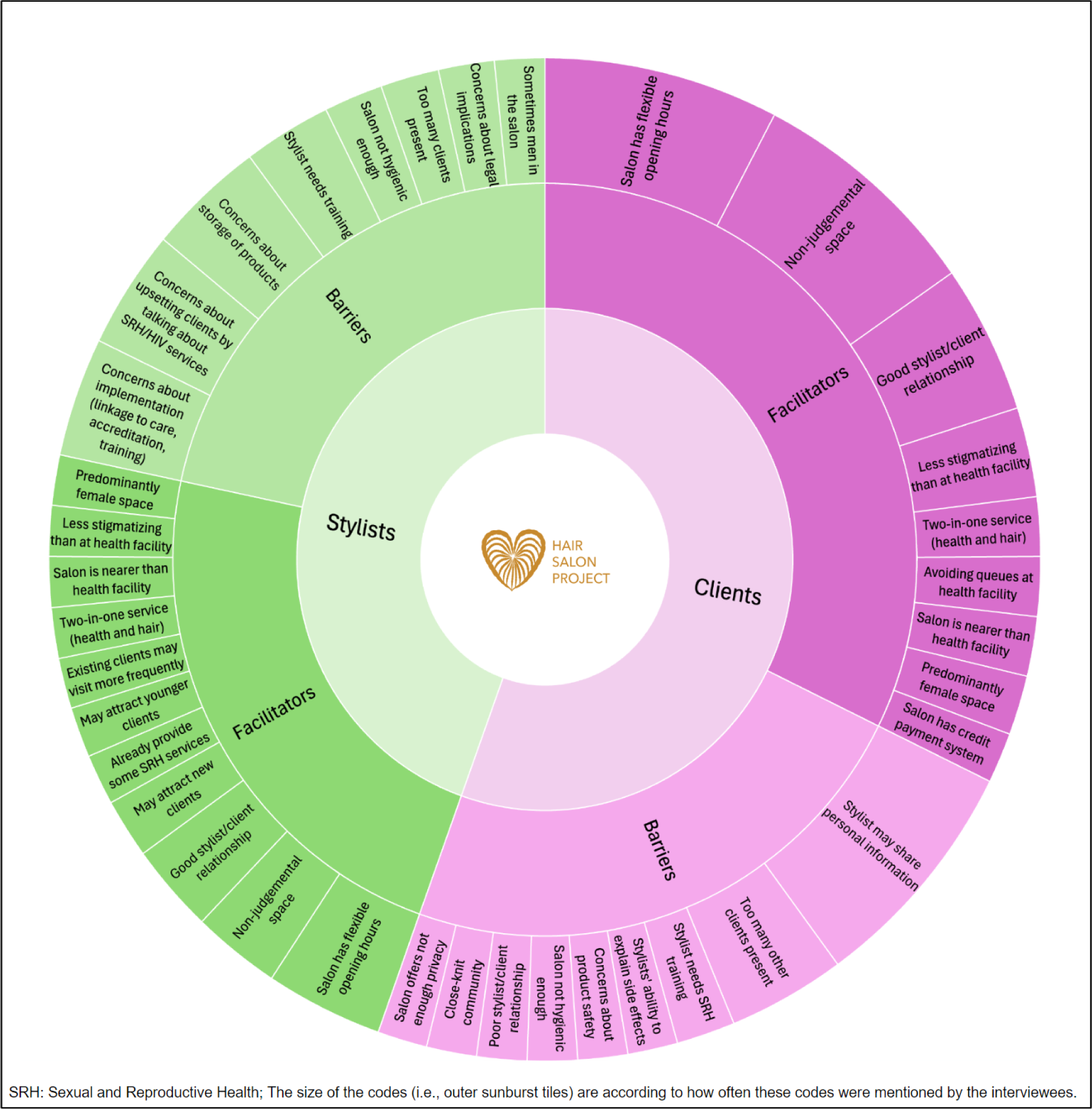
Barriers and Facilitators of offering/receiving HIV/SRH services at the hair salon.

*“I think another benefit can be that at salons, the stylists are able to talk to people so clients can be comfortable talking to them and getting help.”* [P2, client]

*“Yes, I think I’d be comfortable telling my clients that ‘here are some condoms, anyone who needs them can take them as needed”* [P1, stylist]

*“No, I wouldn’t have a problem [providing PEP] because we are also at risk, we could need it*.

*Clients can also be at risk and need it.”* [P7, stylist]

Compared to HIV/SRH services offered at the clinic, many felt less judged at the hair salon:

*“A top challenge for me is that sometimes I don’t want to go to the hospital. Some nurses ask questions that are too judgemental (laughs), yes we are young, and we are already sexually active, but the last thing we need is to be judged”* [P11, client]

*“Sometimes at the clinic, you are not free to talk about [SRH services]. You are being judged. You are talking to a stranger. […] When we are in the queues waiting for services, they specify which queue is for which service and one gets scared to queue on the right line because they believe everyone will see what they are at the clinic for. At the pharmacy too it’s still a challenge, you check who is behind you to see if they will hear that you are there to buy condoms.”* [P3, stylist]

Some interviewees highlighted how integrating HIV services with SRH services as well as general beauty/hair services will facilitate and normalize acceptability of HIV services:

*“ I think since women already go to salons almost every month, it would be easier for them to get contraceptives from there. For example, if she uses monthly contraceptives, she can get two things done in one go: she will get her hair done, and get her contraceptives on the side.*” [P13, client]

Among the low number of stylists and clients who were uncomfortable providing/receiving HIV/SRH services at the hair salon, lack of privacy was the most common concern (Table S6 and S7). For services related to HIV, not feeling confident enough to provide them (among stylists) and preferring a healthcare professional instead (among clients) were the top concerns. Interviewees provided insights in this regard:

*“I think it’s wise for me to attend the workshops about these services so as to have the correct information about them because I’m not a member of the health faculty. What I know is what I hear and what we tell one another. Therefore I think we should have professional training where we would have the correct information to pass on to our clients”* [P7, stylist]

*“I feel like things such as PrEP and HIV are very sensitive. You need someone who is very discrete, and someone with whom you will never have a conflict. I would much rather prefer to get it from the clinic”* [P11, client]

When survey clients were asked which SRH services they would be interested in receiving at the salon in the next six months (multiple answer options possible), HIV counseling was chosen by 14.6%, HIVST by 30.8%, PrEP by 22.1%, and PEP by 14.9% (Table 2). Oral HIVST was chosen similarly among younger and older clients, PrEP by 13.3% of the younger and 28.3% of older, and PEP by 7.0% of the younger and 20.6% of older clients. Family planning services, menstrual products and GBV information provision were chosen by more than a quarter overall, with no major differences between the two client groups. Only 2.9% of clients reported no interest in receiving any services at the salon.

**Table 2.** Sexual and reproductive health services that clients would be interested in if available at their salon in the next six months, stratified by age group.

| Service | Clients |  |  |
| --- | --- | --- | --- |
|  | Overall | 15-24 yrs | 25-35 yrs |
|  | n = 308 | n = 128 | n = 180 |
| Family planning counselling | 176 (57.1) | 65 (50.8) | 111 (61.7) |
| External/male condoms | 75 (24.4) | 27 (21.1) | 48 (26.7) |
| Internal condoms/female condoms | 70 (22.7) | 21 (16.4) | 49 (27.2) |
| Contraceptive pill | 86 (27.9) | 28 (21.9) | 58 (32.2) |
| Emergency contraceptive pill | 90 (29.2) | 39 (30.5) | 51 (28.3) |
| HIV counselling | 45 (14.6) | 16 (12.5) | 29 (16.1) |
| Oral HIV self-testing | 95 (30.8) | 37 (28.9) | 58 (32.2) |
| HIV pre-exposure prophylaxis | 68 (22.1) | 17 (13.3) | 51 (28.3) |
| HIV post-exposure prophylaxis | 46 (14.9) | 9 ( 7.0) | 37 (20.6) |
| Sexually transmitted infection counselling | 54 (17.5) | 16 (12.5) | 38 (21.1) |
| Menstrual counselling | 73 (23.7) | 27 (21.1) | 46 (25.6) |
| Menstrual products | 94 (30.5) | 40 (31.2) | 54 (30.0) |
| Gender-based violence information | 80 (26.0) | 22 (17.2) | 58 (32.2) |
| None | 9 ( 2.9) | 4 ( 3.1) | 5 ( 2.8) |

**Note:** Participants could select more than one answer.

A total of 10.8% of stylists believed that offering HIV/SRH services would negatively impact their business (Table S8):

“*Because some people may not be comfortable, and may feel like when they come to my salon, I assume things about them and their behaviors. […] Some may wonder ‘who is she to tell me about my sexual life?’* “ [P12, stylist]

Conversely, when stylists were asked if these services would have a positive impact on their business, 90.4% agreed, with the main reason being the potential to attract new clients (71.8%, <u>Table S8</u>). This was reflected among the stylist interviewees:

“*Well I think It’s going to bring clients in a sense that they know when they come to my salon they are not going to get services for the hair only […] I feel it’s going to bring me more clients. One would tell the other that when you go to that salon you will also get this and this. So I think it is going to expose me to new clients.”* [P7, stylist]

One stylist also felt that more younger women would be willing to come to her salon as a result:

*“I think there will be more clients like youth. The youth are comfortable where they feel that they are not judged, and where they hear a person talking freely. Sometimes one can experience sexually transmitted diseases but isn’t free to talk, but when they come to the salon they can be able to talk and get help.”* [P3, stylist]

### Feasibility of providing HIV/SRH services at hair salons

86.0% of the hair stylists reported that their hair salon is open seven days a week and 81.5% have a toilet on-site (Table 3). 60.5% mentioned that they are serving three or more clients simultaneously and only 21.0% have an additional confidential space - crucial aspects reflected upon in the interviews:

**Table 3.**
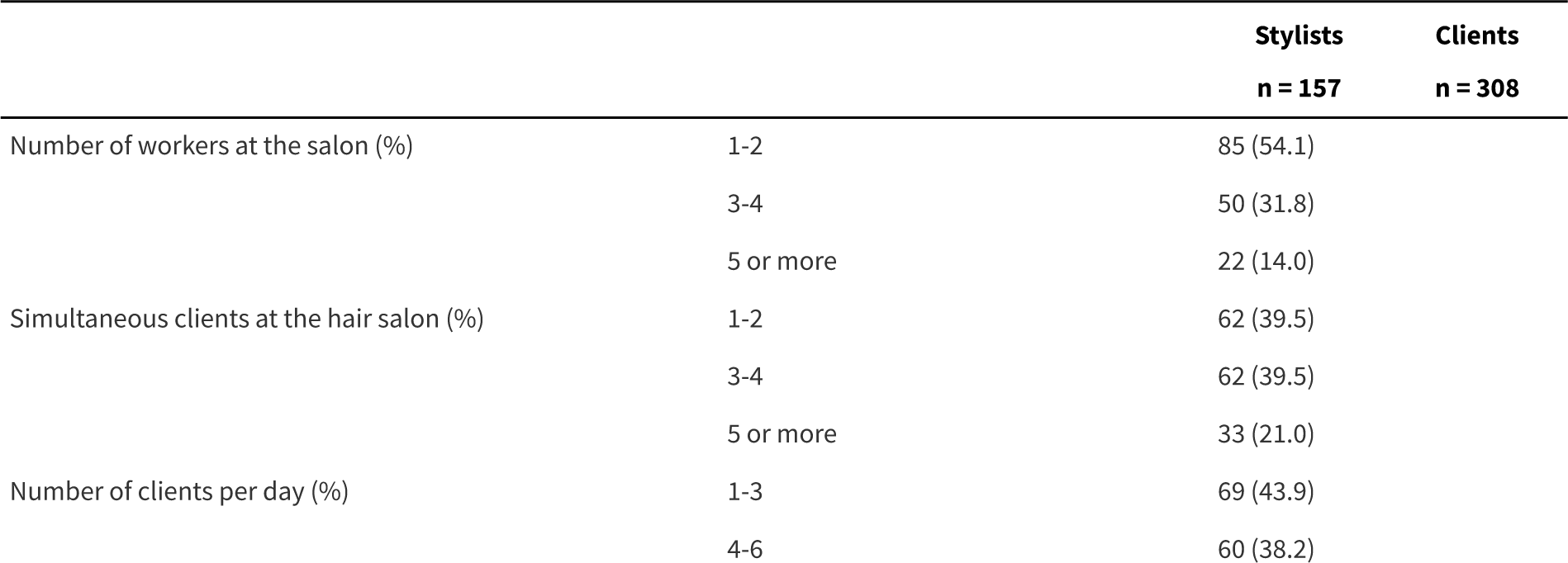
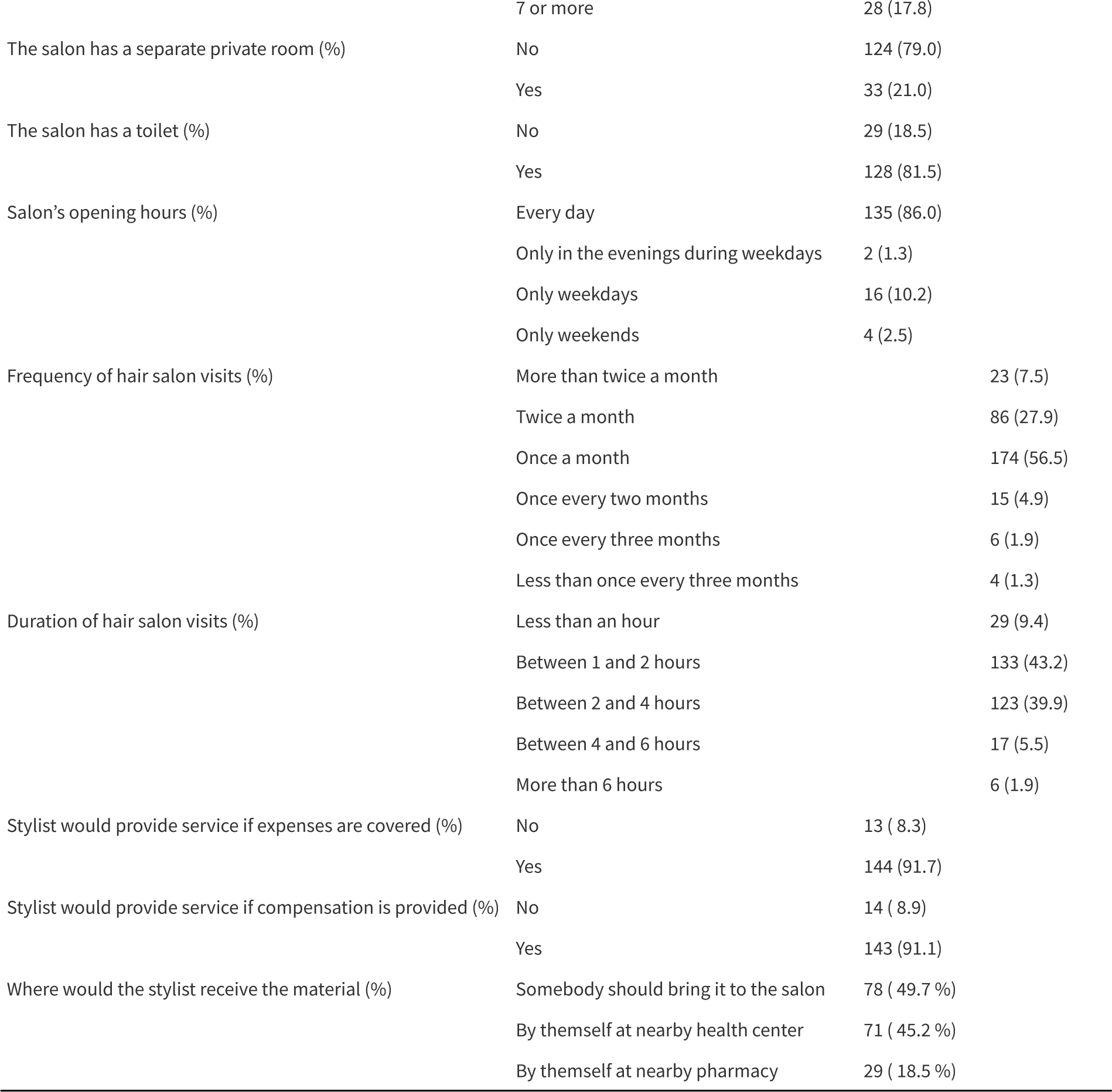
Hair salon characteristics and feasibility measures.

*“You know! Sometimes there’s a random person sitting in the salon who isn’t even there to do her hair. I don’t know, but I think of a cubicle in the same salon space which is enough for a private conversation. If only the client is comfortable moving to the private space to talk, and doesn’t mind that other clients will see her going to that private space. I agree that the salon space is not safe [in terms of privacy] because usually there’s a walk-in service where other people may just drop by and interrupt your conversation. So I think a small cubicle may be a good idea.”* [P1, stylist]

Concerning reimbursement expectations, 91.7% of stylists indicated they would provide HIV/SRH services even if their time is not compensated as long as the costs for the products and supply are covered (Table 3). The most favored supply option for stylists was having someone delivering the HIV/SRV products (49.7%) (Table 3).

56.5% of the clients visit their salon once a month and 35.4% even more frequently (Table 3). For 83.1% of clients, a hair salon visit lasts between 1 and 4 hours. More than 82% of clients reach their hair salon within 30 minutes and 68.1% spend less than 20 Maloti (about $1.14 USD) on travel costs (Table S9). Hair salons seemed slightly more accessible than the nearest healthcare centers in terms of cost and time (Table S9), as mentioned in the interviews too:

*“It could benefit them because walking from my village to the health center is a long distance, so if one goes to a hair salon close by, it wouldn’t take long like walking to the health center.”* [P2, client]

The qualitative data additionally emphasized the salons flexibility in terms of opening hours: *“People usually go to the salons unlike to the clinic or to the pharmacy. Therefore it will be an advantage to my clients to get these services to the salon instead of going to the clinics. I also think the time at the salon is convenient for everyone. At the health centres, we go to on specific times and dates e.g from Monday to Friday maybe 8 am to 4 pm. But at the salon, one goes every time.”* [P7, stylist]

“*is [the emergency contraceptive pill, i.e., plan B] the one that is used within a certain number of hours? […]. It would be better at the salon. At the clinic, I may need to wait in a long queue which would waste more time. But at the salon, I would just go in and get the plan B.”* [P11, client]

## Discussion

This innovative citizen scientist mixed-methods study among 157 hair stylists and 308 of their women clients aged 15-30 assessed the acceptability and feasibility of offering/receiving various HIV/SRH services at hair salons in peri/urban areas across Lesotho. We found high acceptability with more than 83% of stylists and clients being comfortable offering/receiving services including HIV counseling, HIVST, PrEP, PEP, and over 90% of stylists thinking that this would have a positive impact on their business, attracting new and younger clients. Themes such as “safe space” and “non-judgemental” were commonly mentioned. Feasibility was equally high with salons being highly accessible, frequently visited and offering flexible opening hours. However, several concerns were raised, especially related to privacy and stylists’ role and capacity.

The only other published empirical evidence on the same topic from an African setting stems from three studies in South Africa[20-22] and one from Ghana[23]. However, the latter was merely a survey addressing SRH topics among hair stylists during the apprenticeship rather than providing services to their clients[23]. Similar to our study, a survey across 19 hair salons in the townships around Durban, South Africa, found that 98% of stylists felt comfortable offering health education if properly trained and the vast majority was comfortable with clients receiving injectable contraception (98%), PrEP (95%), and HIV testing (87%) provided by a nurse at their salon[21]. Among their women clients, 77% were comfortable receiving PrEP, and 74% HIV testing at the salon. In addition, the team from Durban interviewed >90 clients and stylists, exploring specific services such as contraception and PrEP [20, 22]. They identified similar facilitators to those observed in our study, such as the non-judgemental and high-trust nature of the client-stylist relationship, mutual investment of time at the salon and the high accessibility and conducive environment. Aligning with our findings, the concern most often mentioned was confidentiality. Importantly, the Durban studies are all based on a service model whereby a nurse is attached to the hair salon providing the services. While this offers certain advantages, it may have limitations in terms of sustainability and scalability. Our results suggest that a service model whereby the hair salon stylist is the main service provider reaches similar comfortability levels among both stylists and clients.

We observed substantial disparity between the comfortability levels and the more immediate demand. For example, 88.3% of clients were comfortable receiving PrEP at their hair salon from their stylist. However, when asked which of the 13 assessed HIV/SRH services they would opt for if provided at the hair salon in the coming 6 months, PrEP was chosen only by 22.1%. There are likely several explanations for this. First, not all surveyed clients may have been sexually active or some were already living with HIV or taking PrEP, and therefore should be removed from the denominator. Second, while we found that PrEP knowledge among clients was high (>82%), myths around PrEP were prevalent. Third, not all clients at risk may also perceive themselves at risk, which is an important predictor and mediator for PrEP uptake[45-48]. Self-perceived high HIV risk among surveyed clients was low, especially among the younger clients, comparable to studies from similar settings[49]. Nonetheless, if the 22.1% PrEP demand translated into actual PrEP uptake, this would higher than at post abortion care clinics[50] and comparable to other community initiatives such as community health fairs[51], mobile teen health clinics[52] or pharmacy-based PrEP delivery[53].

The findings of this study are promising, however piloting an HIV/SRH service package at hair salons in Lesotho will provide a clearer understanding. Several recommendations for a pilot were drawn from the results. First, although the hair salon seems to be a trusted and non-judgemental space, privacy concerns were raised in case of actual service provision, especially for HIV services. Only a fifth of the salons reported a separate confidential space and most have several clients in parallel. Some interviewees suggested creating a confidential space within the hair salon (e.g., cubicles) either for confidential counselling or simply for confidential product retrieval. Second, the stylists’ role needs to be clarified and appropriate training ensured to capacitate the stylist. Third, one size may not fit all. The younger clients (15-24 years old) were markedly different in terms of sociodemographic characteristics (marital status, economic status, occupation) than the 25-35 years old clients, with more demand for HIV services among older than younger clients. This may partly be due to the fact that the HIV prevalence, and thus awareness, are higher among the older age group (28-40%) than among the younger age group (5-13%)[5]. However, the majority of clients across both age groups were comfortable with menstrual health and family planning services, signaled high demand for these services, and mentioned their role - besides the beauty/hair services - as facilitators for the HIV services. Fourth, the interaction between HIV/SRH service provision and the hair/beauty business needs to be explored. The majority of stylists thought that HIV/SRH services would positively impact their business by attracting new and potentially younger clients, and thus, agreed to provide them without additional compensation as long as they are appropriately trained, and supply is ensured. Fifth, more than half of the surveyed clients were already active users of different family planning methods. It remains to be seen how many new users of HIV/SRH services can be reached in this space.

The strengths of this study are its nationwide sampling, the in-depth and innovative involvement of citizen scientists and the community, and its rigorous mixed-methods application for a more comprehensive perspective. However, there are limitations. First, due to our sampling approach, the majority of surveyed participants were from peri/urban areas with high levels of education, and thus, the results may not be applicable in rural areas. However, the density of new HIV transmissions in Lesotho is highest in these areas [54], where a future intervention might be most impactful. Second, we lacked more in-depth information about sexual behavior, current and prior PrEP use. Third, we only collected self-reported HIV status indirectly as part of the HIV risk assessment question, resulting in unrealistically low numbers.

## Conclusions

Based on the findings of this nationwide, citizen-science mixed-methods study, offering HIV/SRH services in hair salons in Lesotho seems to be largely acceptable and feasible if confidentiality concerns are mitigated, positioning hair salons as safe community spaces for such services, with many participants feeling less judged at the salon than at clinics. A pilot is warranted to explore how this approach could work in practice.

## Declarations

### Authors’ contributions

PS, TM, TB, DM, JAB, JMB, DJP, MH, KH and AA conceptualized the study and obtained the funding. MC drafted the survey questionnaire over several iterations, reviewed by MK, PS, TM, TB, DM, JAB, JMB, FG, FR, AW, DJP, MH, DC, KH, AA and the Hair SALON Citizen Scientist Working Group. MK and TM translated the questionnaire into Sesotho with support from the Hair SALON Citizen Scientist Working Group. The Hair SALON Citizen Scientist Working Group piloted the questionnaire. MK designed the qualitative part, with input and support from MC, PS, MS, MH, DC, KH, AA, and the Hair SALON Citizen Scientist Working Group, and conducted all interviews. PS and MH led the survey implementation, including social media recruitment and real-time data verification, supervised by MC and AA. AA designed the recruitment and data management and verification tool. MC conducted the quantitative analysis, supervised by AA. MK conducted the qualitative analysis, supervised by DC, KH and AA. MC, MK and AA drafted the first version of the manuscript. All named authors reviewed the draft manuscript, provided input, and approved the final version. Detailed involvement of the Hair SALON Citizen Scientist Working Group provided in <u>Text S2</u>.

### Members of the Hair SALON Citizen Scientist Working Group

Itumeleng Mohale, Thabo Lebakeng, Masia Morena-motšo, Thembekile Mokhosi Mapefane

## Acknowledgements

We are grateful to all clients and stylists who participated in this study. We thank The HUB interns Darlis Dube, Teboho Mokapi and Lehlohonolo Maleke who supported the creation of the data collection tool as well as the data verification and monitoring process.

## Data Availability Statement

We will deposit a de-identified dataset with key data presented in the manuscript on zenodo.org.

## Supporting Information

Supporting Information: Supporting text, figures and tables.

## References

1. Joint United Nations Programme on HIV/AIDS. The urgency of now: AIDS at a crossroads. Geneva 2024 [cited 2024 Aug 21] [Available from: https://www.unaids.org/en/resources/documents/2024/2024-unaids-global-aids-update-adolescent-girls-young-women]

2. Global AIDS. HIV and adolescent girls and young women - Update fact sheet 2023 [cited 2024 Dec 11] [Available from: https://thepath.unaids.org/wp-content/themes/unaids2023/assets/files/thematic_fs_hiv_girls_women.pdf]

3. WHO. Regional Office for Africa. Women’s Health [cited 2024 Dec 12] [Available from: https://www.afro.who.int/health-topics/womens-health]

4. UNAIDS 2024. NAOMI: HIV sub-national estimates. Lesotho [cited 2024 Aug 21] [Available from: https://naomi-spectrum.unaids.org/]

5. Lesotho - Population-Based HIV Impact Assessment. LePHIA 2020: Lesotho Ministry of Health and the Lesotho Bureau of Statistics; 2021 [cited 2024 Dec 11] [Available from: https://phia.icap.columbia.edu/wp-content/uploads/2021/08/53059_14_LePHIA_Summary-sheet_with-coat-of-arms_WEB_v2.pdf]

6. Picchetti V, Stamatakis C, Annor FB, Massetti GM, Hegle J. Association between lifetime sexual violence victimization and selected health conditions and risk behaviors among 13–24-year-olds in Lesotho: Results from the Violence Against Children and Youth Survey (VACS), 2018. Child Abuse Negl. 2022;134:105916.

7. Guttmacher Institute. Lesotho. Unintended pregnancies and abortions. [cited 2024 Aug 21] [Available from: https://www.guttmacher.org/regions/africa/lesotho]

8. Lesotho National Guidelines on the Use of Antiretroviral Therapy 2022 [cited 2024 Dec 11] [Available from: https://hivpreventioncoalition.unaids.org/en/resources/lesotho-national-guidelines-use-antiretroviral-therapy-hiv-prevention-and-treatment-sixth]

9. National Guidelines On The Use Of Antiretroviral Therapy For HIV Prevention And Treatment 2016 [cited 2024 Dec 11] [Available from: https://www.childrenandaids.org/sites/default/files/2017-04/Lesotho_ART-Guidelines_2016.pdf]

10. Dunbar MS, Kripke K, Haberer J, Castor D, Dalal S, Mukoma W, et al. Understanding and measuring uptake and coverage of oral pre-exposure prophylaxis delivery among adolescent girls and young women in sub-Saharan Africa. Sex Health. 2018;15(6):513–21.

11. Palanee-Phillips T. Editorial: Integration of HIV prevention with sexual and reproductive health services. Front Reprod Health. 2023;5:1129881.

12. World Bank Group. Rural population (% of total population). Lesotho [cited 2024 Mar 26] [Available from: https://data.worldbank.org/indicator/SP.RUR.TOTL.ZS?locations=LS]

13. National Health Strategic Plan: Government of Lesotho; 2016 [cited 2024 Dec 12] [Available from: https://www.childrenandaids.org/node/531]

14. Morehead-Gee A, Uskup DK, Omokaro U, Shoptaw S, Harawa NT, Heilemann MV. Relating ‘to her Human Side’: a Grounded Theory analysis of cosmetologists’ and aestheticians’ relationships with clients in Black American beauty salons to inform sexual health interventions. Cult Health Sex. 2023;25(9):1180–97.

15. Roberts-Dobie S, Rasmusson A, Losch ME. The Speak UP! Salon Project: Using Hair Stylists as Lay Health Educators About Unintended Pregnancy. Health Promot Pract. 2018;19(1):31–7.

16. DiVietro S, Beebe R, Clough M, Klein E, Lapidus G, Joseph D. Screening at hair salons: The feasibility of using community resources to screen for intimate partner violence. J Trauma Acute Care Surg. 2016;80(2):223–8.

17. Palmer KNB, Rivers PS, Melton FL, McClelland DJ, Hatcher J, Marrero DG, et al. Health promotion interventions for African Americans delivered in U.S. barbershops and hair salons- a systematic review. BMC Public Health. 2021;21(1):1553.

18. Nadison M, Flamm LJ, Roberts A, Staton T, Wiener L, Locke J, et al. Kaiser Permanente’s Good Health & Great Hair Program: Partnering With Barbershops and Beauty Salons to Advance Health Equity in West Baltimore, Maryland. J Public Health Manag Pract. 2022;28(2):E369–E79.

19. Palmer KNB, Okechukwu A, Mantina NM, Melton FL, Kram NA, Hatcher J, et al. Hair Stylists as Lay Health Workers: Perspectives of Black Women on Salon-Based Health Promotion. Inquiry. 2022;59:469580221093183.

20. Wara NJ, Psaros C, Govere S, Dladla N, Stuckwisch A, Zionts D, et al. Hair salons and stylist-client social relationships as facilitators of community-based contraceptive uptake in KwaZulu-Natal, South Africa: a qualitative analysis. Reprod Health. 2021;18(1):178.

21. Bassett IV, Xu A, Govere S, Thulare H, Frank SC, Psaros C, et al. Contraception and PrEP in South African Hair Salons: Owner, Stylist, and Client Views. J Acquir Immune Defic Syndr. 2018;79(2):e78–e81.

22. Bassett IV, Govere S, Millham L, Frank SC, Dladla N, Thulare H, et al. Contraception, HIV Services, and PrEP in South African Hair Salons: A Qualitative Study of Owner, Stylist, and Client Perspectives. J Community Health. 2019;44(6):1150–9.

23. Owusu SA, Blankson EJ, Abane AM. Sexual and reproductive health education among dressmakers and hairdressers in the Assin South District of Ghana. Afr J Reprod Health. 2011;15(4):109–19.

24. English PB, Richardson MJ, Garzon-Galvis C. From Crowdsourcing to Extreme Citizen Science: Participatory Research for Environmental Health. Annu Rev Public Health. 2018;39:335–50.

25. Todowede O, Lewandowski F, Kotera Y, Ashmore A, Rennick-Egglestone S, Boyd D, et al. Best practice guidelines for citizen science in mental health research: systematic review and evidence synthesis. Front Psychiatry. 2023;14:1175311.

26. Marks L, Laird Y, Trevena H, Smith BJ, Rowbotham S. A Scoping Review of Citizen Science Approaches in Chronic Disease Prevention. Front Public Health. 2022;10:743348.

27. Thomas JA, Trigg J, Morris J, Miller E, Ward PR. Exploring the potential of citizen science for public health through an alcohol advertising case study. Health Promot Int. 2022;37(2).

28. Tan BXH, Chong SY, Ho DWS, Wee YX, Jamal MH, Tan RKJ. Fostering citizen-engaged HIV implementation science. J Int AIDS Soc. 2024;27 Suppl 1(Suppl 1):e26278.

29. Tovin MM, Wormley ME. Systematic Development of Standards for Mixed Methods Reporting in Rehabilitation Health Sciences Research. Phys Ther. 2023;103(11).

30. McNall M, Foster-Fishman PG. Methods of Rapid Evaluation, Assessment, and Appraisal. American Journal of Evaluation. 2007;28(2):151–68.

31. Sekhesa P, Kopeka M, Chiaborelli M, Mohloanyane T, Ballouz T, Menges D, et al. The hair SALON project - A Hair Salon Citizen Scientist Mixed-Methods Study. [Preprint]; 2024 [Available from: https://osf.io/tm26g/]

32. Staniszewska S, Brett J, Simera I, Seers K, Mockford C, Goodlad S, et al. GRIPP2 reporting checklists: tools to improve reporting of patient and public involvement in research. BMJ. 2017;358:j3453.

33. Begnel ER, Escudero J, Mugambi M, Mugwanya K, Kinuthia J, Beima-Sofie K, et al. High pre-exposure prophylaxis awareness and willingness to pay for pre-exposure prophylaxis among young adults in Western Kenya: results from a population-based survey. Int J STD AIDS. 2020;31(5):454–9.

34. Hennegan J, Nansubuga A, Akullo A, Smith C, Schwab KJ. The Menstrual Practices Questionnaire (MPQ): development, elaboration, and implications for future research. Glob Health Action. 2020;13(1):1829402.

35. Hampanda K, Ybarra M, Bull S. Perceptions of health care services and HIV-related health-seeking behavior among Uganda adolescents. AIDS Care. 2014;26(10):1209–17.

36. KoboToolbox [Internet]. KoboToolbox. [cited 2023 Oct 10]. [Available from: https://www.kobotoolbox.org/]

37. Keating P, Murray J, Schenkel K, Merson L, Seale A. Electronic data collection, management and analysis tools used for outbreak response in low- and middle-income countries: a systematic review and stakeholder survey. BMC Public Health. 2021;21(1):1741.

38. Package ‘KoboconnectR’. [cited 2024 Dec 11] [Available from: https://cran.r-project.org/web/packages/KoboconnectR/KoboconnectR.pdf]

39. Chang W, Cheng J, Allaire J, Sievert C, Schloerke B, Xie Y, et al. shiny: Web Application Framework for R. R package version 1.9.1.9000. 2024 [cited 2023 Dec 11] [Available from: https://github.com/rstudio/shiny, https://shiny.posit.co/]

40. Micronutrient Survey Manual & Toolkit. Calculation of sample size for a single cross-sectional cluster survey. [cited 2023 Oct 9] [Available from: https://mnsurvey.nutritionintl.org/categories/13]

41. CDC. Pre-Exposure Prophylaxis (PrEP) Chapter – Evidence-Based Efficacy Criteria 2023 [cited 2024 Oct 11] [Available from: https://www.cdc.gov/hiv/pdf/research/interventionresearch/compendium/prep/PrEP_Chapter_EBI_Criteria.pdf]

42. Sheira LA, Kwena ZA, Charlebois ED, Agot K, Ayieko B, Gandhi M, et al. Testing a social network approach to promote HIV self-testing and linkage to care among fishermen at Lake Victoria: study protocol for the Owete cluster randomized controlled trial. Trials. 2022;23(1):463.

43. Hennink M, Kaiser BN. Sample sizes for saturation in qualitative research: A systematic review of empirical tests. Soc Sci Med. 2022;292:114523.

44. R Core Team. R Foundation for Statistical Computing V, Austria. R: A language and environment for statistical computing. 2021. [Available from: https://www.R-project.org/]

45. Velloza J, Mujugira A, Muwonge T, Boyer J, Nampewo O, Badaru J, et al. A novel “HIV salience and Perception” scale is associated with PrEP dispensing and adherence among adolescent girls and young women in Kampala, Uganda. AIDS Behav. 2023;27(1):279–89.

46. Hill LM, Maseko B, Chagomerana M, Hosseinipour MC, Bekker LG, Pettifor A, et al. HIV risk, risk perception, and PrEP interest among adolescent girls and young women in Lilongwe, Malawi: operationalizing the PrEP cascade. J Int AIDS Soc. 2020;23 Suppl 3(Suppl 3):e25502.

47. Karletsos D, Greenbaum CR, Kobayashi E, McConnell M. Willingness to use PrEP among female university students in Lesotho. PLoS One. 2020;15(3):e0230565.

48. Hampanda K, Bolt M, Nayame L, Hamoonga T, Sehrt M, Thorne J, Harrison M, Pintye J, Amstutz A, Abuogi L, Mweemba O. HIV Risk and Intention to use PrEP among Sexually Active Female University Students in Zambia: A Cross-Sectional Survey to Understand Influential Factors. [Preprint]; 2024 [Available from: https://www.medrxiv.org/content/10.1101/2024.12.12.24318948v1]

49. Heck CJ, Reed DM, Okal J, Chipeta E, Mbizvo M, Mathur S. Examining concordance of sexual-related factors and PrEP eligibility with HIV risk perception among adolescent girls and young women: cross-sectional insights from DREAMS sites in Kenya, Malawi, and Zambia. BMC Public Health. 2024;24(1):2793.

50. Zia Y, Etyang L, Nyerere B, Nyamwaro C, Mogaka F, Mwangi M, et al. Structural influences on delivery and use of oral HIV PrEP among adolescent girls and young women seeking post abortion care in Kenya. EClinicalMedicine. 2024;68:102416.

51. Koss CA, Charlebois ED, Ayieko J, Kwarisiima D, Kabami J, Balzer LB, et al. Uptake, engagement, and adherence to pre-exposure prophylaxis offered after population HIV testing in rural Kenya and Uganda: 72-week interim analysis of observational data from the SEARCH study. Lancet HIV. 2020;7(4):e249–e61.

52. Rousseau E, Bekker LG, Julies RF, Celum C, Morton J, Johnson R, et al. A community-based mobile clinic model delivering PrEP for HIV prevention to adolescent girls and young women in Cape Town, South Africa. BMC Health Serv Res. 2021;21(1):888.

53. Vera M, Bukusi E, Achieng P, Aketch H, Araka E, Baeten JM, et al. “Pharmacies are Everywhere, and You can get it at any Time”: Experiences With Pharmacy-Based PrEP Delivery Among Adolescent Girls and Young Women in Kisumu, Kenya. J Int Assoc Provid AIDS Care. 2023;22:23259582231215882.

54. Low A, Thin K, Davia S, Mantell J, Koto M, McCracken S, et al. Correlates of HIV infection in adolescent girls and young women in Lesotho: results from a population-based survey. Lancet HIV. 2019;6(9):e613–e22.

