## Supporting Information: Supporting text, figures and tables. for "Hair salons as a promising space to provide HIV and sexual and reproductive health services for young women in Lesotho: A citizen scientist mixed-methods study"


Text S1. Reporting according to the Mixed Method Reporting in Rehabilitation & Health Services (MMR-RHS) checklist1. 2
Text S2. Description of the role of community and citizen scientist involvement, based on the Guidance for Reporting Involvement of Patients and the Public (GRIPP2) checklist short form1. 4
Text S3. HIV/Sexual and Reproductive health(SRH) services evaluated in the survey 6
Text S4. Questionnaire for stylists. 7
Text S5. Questionnaire for clients. 28
Text S6. Interview guides. 50
Figure S1. Overview of survey steps and procedures 56
Figure S2. Participants flowchart. 57
Figure S3. Map of the participating 157 hair salons/stylists 58
Table S1. Interviewed stylists and clients' baseline characteristics. 59
Table S2. PrEP, HIVST, Family planning and Menstrual health. 60
Table S3. Comfortability of stylists offering sexual and reproductive health services. 62
Table S4. Comfortability of clients receiving sexual and reproductive health services. 63
Table S5. Barriers and facilitators of offering/receiving HIV/SRH services at hair salons. Qualitative analysis matrix. 64
Table S6. Reasons for stylists' discomfort in offering sexual and reproductive health services. 71
Table S7. Reasons for clients' discomfort in receiving sexual and reproductive health services. 72
Table S8. Impact stylists report believing offering SRH services could have on their business. 73
Table S9. Accessibility of hair salons in terms of both cost and time. 74

Text S1. Reporting according to the Mixed Method Reporting in Rehabilitation & Health Services (MMR-RHS) checklist[1].

| | Yes/No; location |
|---|---|
| **Title** | |
| Concisely describes the topic of the study identifying the study as mixed methods | Yes; title page. |
| **Abstract** | |
| Summarizes key elements using journal specific abstract format; For example:<br>Introduction, Methods, Results, Discussion, and Significance/potential impact to rehabilitation and/or societal health | Yes; abstract page 3. |
| **Introduction** | |
| Includes literature review on the topic of interest (quantitative, qualitative, and mixed) | Yes; introduction page 4 and Discussion page 17. |
| Identifies gap that justifies the need for mixed methods approach | Yes, introduction page 5 and methods page 6. |
| Clearly states overarching goal of the study that supports a mixed methods approach | Yes, introduction pages 4-5. |
| States the rationale for using mixed methods research | Yes, methods pages 4-5. |
| Clearly identifies discrete aim(s) for qualitative and quantitative components. Aims align with corresponding component methods | Yes, methods pages 5-6. |
| Provides statement of significance and potential impact | Yes, introduction page 5. |
| **Methods** | |
| **Design** — Clearly describes the mixed methods design (exploratory sequential, explanatory sequential, concurrent, etc.) used to accomplish the overarching goal of the project:<br>● Emphasis noted (i.e., Sequential QUAL--> quan or QUAN--> qual; Concurrent QUAL + QUAN)<br>● Visual display of overall design highlighting integration (e.g., model, flow chart, figure) | Yes, methods page 5. |
| Describes and supports the qualitative and quantitative methodologies (phenomenology, randomized control trial) used to accomplish the discrete aim(s) of the project | Yes, methods pages 5-7. |

| | |
|---|---|
| States researcher(s) background and contributions to project (e.g., content or methods expertise, relationships to participants) | Yes, methods pages 7-8 and Authors' contributions page 20. |
| Identifies setting (e.g., hospital system, geographical location) | Yes, methods pages 5 and 7. |
| **Data collection** - Clearly describes and supports the following:<br>● Pilot study (if applicable)<br>● Instrumentation (validity, reliability)<br>● Implementation matrix (e.g., data source, timeline, type, anticipated outcomes) | Yes, methods pages 5, 7 and 8. |
| **Data analysis** - Clearly states and describes analysis procedures for:<br>● Qualitative<br>● Quantitative<br>● Mixed Methods (integration) | Yes, methods pages 5-7. |
| **Methodological Rigor** — Clearly describes steps taken to establish rigor:<br>● Qualitative (e.g., credibility, dependability, confirmability, transferability)<br>● Quantitative (e.g., validity, reliability, generalizability)<br>● Mixed Methods (validity or legitimacy) | Yes, methods pages 5-8. |
| **Results/Findings** | |
| Clearly presents findings for study components:<br>● Qualitative (includes data exemplars)<br>● Quantitative<br>● Mixed Methods-Provides integrated findings/overall study results (e.g., joint display) | Yes, results pages 8-17. |
| **Discussion** | |
| Incorporates discussion on implications of integrated findings | Yes, discussion page 17. |
| Provides synthesis and interpretation of findings in the context of existing literature and theoretical/conceptual framework | Yes, discussion page 17-18. |
| Includes subsection of limitations | Yes, discussion page 17-19. |

[1] Tovin MM, Wormley ME. Systematic Development of Standards for Mixed Methods Reporting in Rehabilitation Health Sciences Research. Phys Ther. 2023 Nov 4;103(11):pzad084.

Text S2. Description of the role of community and citizen scientist involvement, based on the Guidance for Reporting Involvement of Patients and the Public (GRIPP2) checklist short form[1].

| | |
|---|---|
| **Aim of community and citizen scientist involvement in this study** | 1. To co-design the most critical aspects of this mixed-methods study with stylists and clients<br>2. To draw on the experience and expertise of stylists and clients<br>3. To render the research accessible to the affected community |
| **Methods used for community and citizen scientist involvement in this study** | ➢ We set up a core citizen scientist group, the Hair SALON Citizen Scientist Working Group (CSWG) consisting of 4 members, and agreed on tasks, responsibilities and reimbursement by signed agreements<br>➢ The CSWG received several trainings on i) the survey content, ii) qualitative research (esp. interviewing techniques) and iii) creative content creation<br>➢ We conducted a series of workshops. First, a workshop (3 days) to finalize and pilot the survey questionnaire. Second, a workshop (2 days) to finalize and pilot the interview guides. Third, a workshop (2 half days) to discuss the results. And fourth, a workshop (4 days) to create several creative result dissemination outputs for community engagement.<br>➢ All workshops were facilitated at The HUB in Morija, face to face, by a trained facilitator<br>➢ Besides the CSWG, for the survey we engaged 157 stylists across the country, via social media to recruit their clients.<br>➢ The 157 stylists received 100 LSL (5.8 USD) via M-PESA per successfully submitted client questionnaire to reimburse their time and they received a citizen scientist certificate for participation. The clients received a token of appreciation of the value of a basic hair style via M-PESA upon completion of the questionnaire (cornrows; 50 LSL, ca. 2.9 USD). |
| **Result of community and citizen scientist involvement in this study** | ➢ During the first two workshop series they adapted our research tools, based on their concrete experience concerning the hair salon setting<br>➢ For the third workshop, they produced a document of recommendations for a future pilot, based on the results, which we incorporated into the discussion section of the manuscript.<br>➢ The fourth workshop resulted in 6 educational short films for tiktok/facebook/whatsapp, in the local language, featuring the CSWG members as actors:<br>○ 1: Reusable sanitary pads (to address a few questions from the survey about reusable pads, e.g. bad odour)<br>○ 2. PrEP and PEP (to educate people about the difference between PEP and PrEP)<br>○ 3. Gender-based violence (to ask people to take action if they see others being abused)<br>○ 4. HIVST (the accuracy of HIV self-testing)<br>○ 5. Morning-after pill ( education on the timeframe of the morning-after pills and advice that one should take them as soon as possible) |

| | |
|---|---|
| | ○ 6. Birth control pill (effectiveness and effects of the pill)<br>➢ https://www.facebook.com/thehubatmorija/posts/pfbid02QhzAP2MXr6Em1Vc6YuApwcTojcEsuwfRbGW2fBhPqHXnZAFV9x4BQ7eLCVFUCL7zl?locale=en_GB<br>➢ Some of the CSWG were actively involved in the video documentary and provided extensive input: https://thehubatmorija.co.ls/2024/10/21/exploring-hair-salons-as-potential-venues-for-the-promotion-of-sexual-and-reproductive-health-services-in-lesotho/<br>➢ The successful remote engagement of citizen scientists across the country led to a fast recruitment of the needed number of clients in under 4 months |
| **Discussion on the extent to which community and citizen scientist involvement influenced this study overall** | ➢ Citizen scientists enriched the study in several ways<br>➢ They led to a successful and fast recruitment of clients<br>➢ The CSWGs' ideas improved our research tools and their recommendations, based on the results discussion, influenced our interpretation of the findings<br>➢ The CSWG created content that was accessible to the local community<br>➢ The CSWG not only provided recommendations for the next step (a pilot), but also took on a more active role in the results dissemination. |
| **Critical perspective on the inclusion of community and citizen scientist involvement in this study** | ➢ Creating a CSWG also comes with responsibilities and expectations, for example to remain long-term involved. These expectations were not all formalized in the initial agreement and had to be incorporated.<br>➢ Investing enough time in training the CSWG was key. Once capacitated, motivation was high to be even more involved (e.g. as interviewers for a next qualitative study) |

[1] Staniszewska S, Brett J, Simera I, Seers K, Mockford C, Goodlad S, et al. GRIPP2 reporting checklists: tools to improve reporting of patient and public involvement in research. BMJ 2017;358:j3453.

Text S3. HIV/Sexual and Reproductive health(SRH) services evaluated in the survey

1. **Counselling/Info and Referral:** Counselling and information about how HIV can spread and steps that can be taken to protect yourself and your partner. Referral to a nearby health facility to obtain more information.
2. **Screening/Testing:** Offer oral HIV self-testing, and referral after test outcome.
3. **PrEP (Pre-exposure prophylaxis):** Offer oral HIV self-testing, and PrEP.
4. **PEP (Post-exposure prophylaxis):** Offer oral HIV self-testing, and PEP.
5. **FP counselling/Info and Referral:** Counselling and information that aims to support young women in making informed decisions about having children or not, how many, and when. Referral to a nearby health facility to obtain family planning methods.
6. **STI:** Counselling/Info and Referral: Counselling and information about how STIs can spread and steps that can be taken to protect yourself and your partner. Referral to a nearby health facility to obtain more information and testing possibilities.
7. **Male/external condom distribution**: distribute male/external condoms and explain their use.
8. **Female/internal condom distribution:** Distribute female/internal condoms and explain their use.
9. **Oral birth control pill:** Distribute oral birth control pills and explain their use.
10. **Emergency contraception:** distribute emergency contraception and explain its use.
11. **Information and Referral:** Information/flyer about GBV. Referral to an appropriate service to obtain more information and help.
12. **Counselling/Info and Referral:** Counselling and information about how to handle your period better and hygiene around menstrual health. Referral to a nearby health facility or pharmacy to obtain more information and menstrual products.
13. **Menstrual product distribution:** Offer menstrual products (mainly reusable cloth sanitary pads).

Text S4. Questionnaire for stylists.

### Stylist Survey

#### Before we start

We want to understand if it is a good idea to offer sexual and reproductive health services at hair salons in Lesotho. We want to hear your opinion about this.

You can change the language to Sesotho if you want (click on "English" at the top).

#### Before we start

* I understand that my participation in this survey is voluntary and that I may withdraw my consent at any time.

◯ OK

* I understand that my answers will be kept confidential and will only be used for research purposes.

◯ OK

* I understand that I will be remunerated M100 (via M-Pesa) for each client that I recruit. I can recruit maximum 3 clients. So I can receive maximum M300.

◯ OK

#### About yourself

**Please enter your first name**

**Please enter your surname (last name)**

**How old are you?**

**How would you describe your gender?**

◯ Woman

◯ Man

◯ I prefer to self-describe, below

**Other, please describe your gender**

**What is your Whatsapp number?**

______________________________

#### About your hair salon

**Are you the owner of the hair salon?**

- ◯ Yes
- ◯ No

**In which district is your hair salon located?**

- ◯ Butha Buthe
- ◯ Leribe
- ◯ Berea
- ◯ Maseru
- ◯ Mafeteng
- ◯ Mohale's Hoek
- ◯ Quthing
- ◯ Qacha's Nek
- ◯ Mokhotlong
- ◯ Thaba-Tseka

#### About yourself, more information

Remember, we are only interested in your information for research purposes and not to identify you personally. We will not share your personal information with anyone.

**For how many years have you worked as a hair stylist?**

- ◯ 0-1 year
- ◯ 1-3 years
- ◯ 3-6 years
- ◯ more than 6 years

**What is the highest level of school/degree you have completed?**

- ◯ None
- ◯ Primary
- ◯ High School
- ◯ Vocational School
- ◯ Tertiary
- ◯ Prefer not to answer

**Do you identify with any of the following religions?**

- ◯ Christianity
- ◯ Islam
- ◯ Other
- ◯ None
- ◯ Prefer not to answer

**Other, please describe**

___

#### Now, let's talk about family planning methods

**How much do you agree with the following statement: If I wanted contraception/family planning, I know of a place where I can obtain it.**

- ◯ Strongly agree
- ◯ Agree
- ◯ Neither agree nor disagree
- ◯ Disagree
- ◯ Strongly disagree

**From where have you ever gotten information about contraception/family planning methods?**

*Select one or several answers*

- ☐ Health center/hospital
- ☐ From friends
- ☐ From family
- ☐ At school
- ☐ Internet/social media
- ☐ Never received any information
- ☐ Other place

**Other, please describe the location**

______________________________

**Are you (or your sexual partner/s) CURRENTLY using any contraception/family planning method?**

- ◯ Yes
- ◯ No
- ◯ Prefer not to answer

**Which method of contraception/family planning are you (or your sexual partner/s) currently using?**

*Select one or several answers*

- ☐ Male/external condoms
- ☐ Female/internal condoms
- ☐ Oral birth control pill
- ☐ Injectable (e.g., Depo-Provera)
- ☐ Implant (e.g., Implanon)
- ☐ Intrauterine device (IUD)
- ☐ Vaginal Ring
- ☐ Emergency contraception
- ☐ Rhythm (fertility awareness)
- ☐ Withdrawal
- ☐ Other

**Other, please describe your contraception/family planning method**

______________________________

**From where do you obtain your current contraception/family planning method?**

*Select one or several answers*

- ☐ Government health center/hospital
- ☐ Private health center/hospital
- ☐ Family planning clinic (e.g. LPPA)
- ☐ Pharmacy
- ☐ Community health worker/fieldworker
- ☐ Friend/relative
- ☐ Other

**Other, please describe where you obtain your current contraception/family planning method**

______________________________

#### Now, let's talk about menstrual health products. These are products that help you to handle your period.

**Have you ever had a period/menstruation?**

- ◯ Yes
- ◯ No
- ◯ I prefer not to answer

##### » If you ever had a period/menstruation

**During your last menstrual period, what were all the materials you used to catch/absorb your menstruation when you were AT HOME?**

*Select one or several answers*

- ☐ Disposable sanitary pad
- ☐ Reusable (cloth) sanitary pad
- ☐ Tampon
- ☐ Cotton wool
- ☐ Toilet paper
- ☐ Cloth towel
- ☐ Natural material (e.g., leaves, sand, grass)
- ☐ Period underwear
- ☐ Underwear alone
- ☐ Mattress or foam
- ☐ Menstrual cup
- ☐ Other

**Other, please describe the product**

________________________________________

**During your last menstrual period, what were all the materials you used to catch/absorb your menstruation when you were AWAY FROM HOME (e.g. at school/at work)?**

*Select one or several answers*

- [ ] Disposable sanitary pad
- [ ] Reusable (cloth) sanitary pad
- [ ] Tampon
- [ ] Cotton wool
- [ ] Toilet paper
- [ ] Cloth towel
- [ ] Natural material (e.g., leaves, sand, grass)
- [ ] Period underwear
- [ ] Underwear alone
- [ ] Mattress or foam
- [ ] Menstrual cup
- [ ] Other

**Other, please describe the product**

______________________________

**Would you like to learn more about reusable, washable, cloth sanitary pads for your period?**

- ◯ Yes
- ◯ No
- ◯ I don't know what 'reusable, washable cloth sanitary pads' are

**Why not?**

*Select one or several answers*

- [ ] I think they are uncomfortable
- [ ] I think they are not hygienic
- [ ] I think they are not safe/absorbing enough
- [ ] I think they are expensive
- [ ] Other

**Other, please describe why not**

______________________________

**What do you think about the reusable, washable, cloth sanitary pads?**

*Select one or several answers*

- ☐ I think they are comfortable
- ☐ I think they are NOT comfortable
- ☐ I think they are hygienic
- ☐ I think they are NOT hygienic enough
- ☐ I think they are safe/absorbing enough
- ☐ I think they are NOT safe/absorbing enough
- ☐ I think they are cheap
- ☐ I think they are expensive
- ☐ Other

**Other, please describe**

---

**Have you ever missed any school or work day due to your period/menstruation?**

- ○ Yes
- ○ No
- ○ I prefer not to answer

**How often in the past year have you missed school/work due to your period/menstruation?**

- ○ 1-2 times
- ○ 3-4 times
- ○ 5-6 times
- ○ More than 6 times

**What was the most common reason?**

- ○ Because of period pain/discomfort
- ○ Because I did not have access to menstruation health products
- ○ I prefer not to answer

**How old were you when you had your first period/menstruation?**

- ◯ Below 12 years of age
- ◯ Around 12 years of age
- ◯ Around 13 years of age
- ◯ Around 14 years of age
- ◯ Around 15 years of age
- ◯ Older than 15 years of age

#### Now, let's talk about HIV testing and Pre-Exposure Prophylaxis (PrEP) against HIV. PrEP is a pill you can take to prevent getting HIV.

**Have you ever heard of a pill, called pre-exposure prophylaxis (PrEP), to prevent getting HIV?**

- ◯ Yes
- ◯ No

How much do you agree with the following statements?

---

**PrEP is only for people who have many sexual partners**

- ◯ Strongly agree
- ◯ Agree
- ◯ Neither agree nor disagree
- ◯ Disagree
- ◯ Strongly disagree

**PrEP will cause people to have more risky sex**

- ◯ Strongly agree
- ◯ Agree
- ◯ Neither agree nor disagree
- ◯ Disagree
- ◯ Strongly disagree

**Only sex workers need PrEP**

- Strongly agree
- Agree
- Neither agree nor disagree
- Disagree
- Strongly disagree

**Only people with partner(s) living with HIV need PrEP**

- Strongly agree
- Agree
- Neither agree nor disagree
- Disagree
- Strongly disagree

**Instead of taking PrEP, people should just pick their partners carefully**

- Strongly agree
- Agree
- Neither agree nor disagree
- Disagree
- Strongly disagree

**Taking PrEP once provides lifelong protection against HIV**

- Strongly agree
- Agree
- Neither agree nor disagree
- Disagree
- Strongly disagree

**Taking PrEP prevents pregnancy**

- Strongly agree
- Agree
- Neither agree nor disagree
- Disagree
- Strongly disagree

**Taking PrEP protects me against other sexually transmitted infections such as syphilis**

- Strongly agree
- Agree
- Neither agree nor disagree
- Disagree
- Strongly disagree

**Have you ever heard of the HIV self-test, a test that you can do yourself at home using a swab in your mouth to test for HIV?**

- Yes
- No

**What do you think your risk of getting HIV in the next year is?**

- No risk at all
- Small risk
- 50/50 (medium) risk
- High risk
- Very high risk
- I have been diagnosed with HIV
- I don't know
- I prefer not to answer

#### Now, let's talk about different Sexual and Reproductive Health (SRH) services. These are services to help you with your family planning needs, sexual well-being, and sexually transmitted diseases.

When answering the following questions, assume that you will receive appropriate training and all the material for free

How much do you agree with the following statements?

**I am comfortable offering family planning/contraception counselling at my hair salon**

*This means giving information to support young women in making decisions about having children or not, how many, and when, and where to obtain family planning methods.*

- ◯ Strongly agree
- ◯ Agree
- ◯ Neither agree nor disagree
- ◯ Disagree
- ◯ Strongly disagree

**Why do you disagree?**

*Select one or several answers*

- ☐ I think the hair salon is not a place to share such confidential information/issues
- ☐ I think I am not capable enough even if I receive training
- ☐ Religious beliefs
- ☐ Other reason

**Other reason, please describe**

________________________________________

**I am comfortable offering male/external condoms (for free) in my hair salon.**

*Male/external condoms are like protective shields that are worn on the penis during sex. They protect agains pregnancy and sexually transmitted infections.*

- ◯ Strongly agree
- ◯ Agree
- ◯ Neither agree nor disagree
- ◯ Disagree
- ◯ Strongly disagree

**Why do you disagree?**

*Select one or several answers*

- ☐ I think the hair salon is not a place to share such confidential information/issues
- ☐ I think I am not capable enough even if I receive training
- ☐ Religious beliefs
- ☐ Other reason

**Other reason, please describe**

________________________________________

**I am comfortable offering female/internal condoms (for free) in my hair salon.**

*Female/internal condoms are like protective shields that are inserted into the vagina during sex. They protect against pregnancy and sexually transmitted infections.*

- ◯ Strongly agree
- ◯ Agree
- ◯ Neither agree nor disagree
- ◯ Disagree
- ◯ Strongly disagree

**Why do you disagree?**

*Select one or several answers*

- ☐ I think the hair salon is not a place to share such confidential information/issues
- ☐ I think I am not capable enough even if I receive training
- ☐ Religious beliefs
- ☐ Other reason

**Other reason, please describe**

---

**I am comfortable offering the oral birth control pill (for free) in my hair salon.**

*Oral birth control pills are a type of medicine that women take daily by mouth to prevent pregnancy.*

- ◯ Strongly agree
- ◯ Agree
- ◯ Neither agree nor disagree
- ◯ Disagree
- ◯ Strongly disagree

**Why do you disagree?**

*Select one or several answers*

- ☐ I think the hair salon is not a place to share such confidential information/issues
- ☐ I think I am not capable enough even if I receive training
- ☐ Religious beliefs
- ☐ Other reason

**Other reason, please describe**

---

**I am comfortable offering the emergency contraception pill (for free) in my hair salon.**

*Emergency contraception pills are a type of medicine that women can take right after unprotected sex to prevent a pregnancy.*

- ◯ Strongly agree
- ◯ Agree
- ◯ Neither agree nor disagree
- ◯ Disagree
- ◯ Strongly disagree

**Why do you disagree?**

*Select one or several answers*

- ☐ I think the hair salon is not a place to share such confidential information/issues
- ☐ I think I am not capable enough even if I receive training
- ☐ Religious beliefs
- ☐ Other reason

**Other reason, please describe**

___

**I am comfortable offering HIV counselling in my hair salon.**

*This means giving information about how HIV can spread and steps that can be taken to protect oneself and others, as well as information about where to obtain more information and testing possibilities.*

- ◯ Strongly agree
- ◯ Agree
- ◯ Neither agree nor disagree
- ◯ Disagree
- ◯ Strongly disagree

**Why do you disagree?**

*Select one or several answers*

- ☐ I think the hair salon is not a place to share such confidential information/issues
- ☐ I think I am not capable enough even if I receive training
- ☐ Religious beliefs
- ☐ Other reason

**Other reason, please describe**

___

**I am comfortable offering a test kit for oral HIV self-testing (for free) in my hair salon.**

*A test kit for oral HIV self-testing includes a swab that allows you to collect fluid yourself from inside your mouth to test for HIV. It is designed to allow you to take the HIV test in private and anonymously.*

- ◯ Strongly agree
- ◯ Agree
- ◯ Neither agree nor disagree
- ◯ Disagree
- ◯ Strongly disagree

**Why do you disagree?**

*Select one or several answers*

- ☐ I think the hair salon is not a place to share such confidential information/issues
- ☐ I think I am not capable enough even if I receive training
- ☐ Religious beliefs
- ☐ Other reason

**Other reason, please describe**

---

**I am comfortable offering pre-exposure prophylaxis (PrEP), for free, in my hair salon.**

*PrEP is a pill that people can take to prevent getting HIV if someone is exposed to it.*

- ◯ Strongly agree
- ◯ Agree
- ◯ Neither agree nor disagree
- ◯ Disagree
- ◯ Strongly disagree

**Why do you disagree?**

*Select one or several answers*

- ☐ I think the hair salon is not a place to share such confidential information/issues
- ☐ I think I am not capable enough even if I receive training
- ☐ Religious beliefs
- ☐ Other reason

**Other reason, please describe**

---

**I am comfortable offering post-exposure prophylaxis (PEP), for free, in my hair salon.**

*PEP is a pill that people can take if they think they have been exposed to HIV. It's like an emergency treatment that helps prevent the virus from spreading in the body.*

- ◯ Strongly agree
- ◯ Agree
- ◯ Neither agree nor disagree
- ◯ Disagree
- ◯ Strongly disagree

**Why do you disagree?**

*Select one or several answers*

- ☐ I think the hair salon is not a place to share such confidential information/issues
- ☐ I think I am not capable enough even if I receive training
- ☐ Religious beliefs
- ☐ Other reason

**Other reason, please describe**

---

**I am comfortable offering counselling about Sexually Transmitted Infections (STIs/STDs), for free, in my hair salon.**

*This means giving information about how STIs can spread and steps that can be taken to protect yourself. And information about where to obtain more information and testing possibilities.*

- ◯ Strongly agree
- ◯ Agree
- ◯ Neither agree nor disagree
- ◯ Disagree
- ◯ Strongly disagree

**Why do you disagree?**

*Select one or several answers*

- ☐ I think the hair salon is not a place to share such confidential information/issues
- ☐ I think I am not capable enough even if I receive training
- ☐ Religious beliefs
- ☐ Other reason

**Other reason, please describe**

---

**I am comfortable offering counselling about menstrual health in my hair salon.**

*This means offering information about how to handle the period and hygiene around menstrual health. And information about where to obtain menstrual products.*

- ◯ Strongly agree
- ◯ Agree
- ◯ Neither agree nor disagree
- ◯ Disagree
- ◯ Strongly disagree

**Why do you disagree?**

*Select one or several answers*

- ☐ I think the hair salon is not a place to share such confidential information/issues
- ☐ I think I am not capable enough even if I receive training
- ☐ Religious beliefs
- ☐ Other reason

**Other reason, please describe**

________________________________________

**I am comfortable offering menstrual products (for example reusable cloth sanitary pads), for free, in my hair salon.**

*Menstrual products such as the reusable cloth sanitary pads absorb the menstrual flow and help you to stay clean and comfortable during your period.*

- ◯ Strongly agree
- ◯ Agree
- ◯ Neither agree nor disagree
- ◯ Disagree
- ◯ Strongly disagree

**Why do you disagree?**

*Select one or several answers*

- ☐ I think the hair salon is not a place to share such confidential information/issues
- ☐ I think I am not capable enough even if I receive training
- ☐ Religious beliefs
- ☐ Other reason

**Other reason, please describe**

________________________________________

**I am comfortable offering gender-based violence (GBV) information in my hair salon.**

*This means providing a space for people who have experienced violence or abuse based on their gender to talk about it. And information about where to get further help.*

- ◯ Strongly agree
- ◯ Agree
- ◯ Neither agree nor disagree
- ◯ Disagree
- ◯ Strongly disagree

**Why do you disagree?**

*Select one or several answers*

- ☐ I think the hair salon is not a place to share such confidential information/issues
- ☐ I think I am not capable enough even if I receive training
- ☐ Religious beliefs
- ☐ Other reason

**Other reason, please describe**

________________________________________

#### Now, let's talk about your hair salon business and the possible impact of offering Sexual and Reproductive Health (SRH) services on your business.

**How much do you agree with the following statement: Providing any of these services would have a POSITIVE impact on my business.**

- ◯ Strongly agree
- ◯ Agree
- ◯ Neither agree nor disagree
- ◯ Disagree
- ◯ Strongly disagree

**Why?**

*Select one or several answers*

- ☐ May attract new clients
- ☐ Usual clients may come more frequently
- ☐ Other reason

**Other, please describe**

________________________________________

**How much do you agree with the following statement: Providing any of these services would have a NEGATIVE impact on my business.**

- ◯ Strongly agree
- ◯ Agree
- ◯ Neither agree nor disagree
- ◯ Disagree
- ◯ Strongly disagree

**Why?**

*Select one or several answers*

- ☐ I may lose working hours
- ☐ Clients may not come to my hair salon because they dislike the idea of being offered SRH services
- ☐ Other reason

**Other, please describe**

______________________________

**What are the opening hours of your hair salon?**

- ◯ Everyday, even weekends, the whole day
- ◯ Only during weekdays, the whole day
- ◯ Only on weekends
- ◯ Only in the evenings during weekdays

**Does your hair salon have a separate private room to talk to clients confidentially?**

- ◯ Yes
- ◯ No

**Does your hair salon offer a toilet for clients?**

- ◯ Yes
- ◯ No

**How many people work in your hair salon, including you?**

- ◯ 1
- ◯ 2
- ◯ 3
- ◯ 4
- ◯ 5
- ◯ 6
- ◯ more than 6

**How many clients are AT THE SAME TIME in your hair salon on a usual working day?**

- ◯ 1
- ◯ 2
- ◯ 3
- ◯ 4
- ◯ 5
- ◯ 6
- ◯ more than 6

**How many clients do you see during a typical working day?**

- ◯ 1
- ◯ 2
- ◯ 3
- ◯ 4
- ◯ 5
- ◯ 6
- ◯ 7
- ◯ 8
- ◯ 9
- ◯ 10
- ◯ more than 10

#### Now, let's talk about the financial support and the supply you would need to provide such services

**How much do you agree with the following statement: I would be willing to offer Sexual and Reproductive Health (SRH) services to my clients if my expenses (material, travel, training) are covered.**

- ◯ Strongly agree
- ◯ Agree
- ◯ Neither agree nor disagree
- ◯ Disagree
- ◯ Strongly disagree

**How much do you agree with the following statement: I would be willing to provide Sexual and Reproductive Health (SRH) services to my clients if - besides covering my expenses (material, travel, training) - I also receive financial compensation for my efforts.**

- ◯ Strongly agree
- ◯ Agree
- ◯ Neither agree nor disagree
- ◯ Disagree
- ◯ Strongly disagree

**How and where would you prefer to receive the material (for example condoms) on a regular basis?**

*Select one or several answers*

- ☐ I would regularly go to the nearby health center/hospital
- ☐ I would regularly go to the nearby pharmacy to pick up the material
- ☐ Someone should regularly come and bring the material

#### Final details

* Once you submit this survey, we will check your answers. This will take a few hours or days.

◯ OK

* Then, please recruit up to 3 clients, let them register on the project homepage (https://hairsalonproject.com/clients/) and let them fill in their questionnaire. Once done, you will receive your reimbursement.

◯ OK

If you have any questions, please call the HUB (+266 5708 7179).

---

Please sign on the next screen. Afterwards, you will be redirected to a website where we explain again all the next steps to follow to get your reimbursement.

---

**Please sign here and submit.**

*Draw your signature with your finger.*

Text S5. Questionnaire for clients.

### Client Survey

#### Before we start

We want to understand if it is a good idea to offer sexual and reproductive health services at hair salons in Lesotho. We want to have your opinion.

You can change the language to Sesotho if you want (click on "English" at the top).

#### Before we start

* I understand that my participation in this survey is voluntary and that I may withdraw my consent at any time.

◯ OK

* I understand that my answers will be kept confidential and will only be used for research purposes.

◯ OK

* I understand that I will receive M50 (via M-Pesa) as a small gift after participating in the survey.

◯ OK

#### About yourself

**Please enter your first name**

______________________________

**Please enter your last name (surname)**

______________________________

**How old are you?**

______________

**How would you describe your gender?**

◯ Woman

◯ Man

◯ I prefer to self-describe, below

Other, please describe your gender

______________________________

**What is your Whatsapp number?**

______________________

#### About your hair salon

**In which district is the hair salon you are visiting?**

- ◯ Butha Buthe
- ◯ Leribe
- ◯ Berea
- ◯ Maseru
- ◯ Mafeteng
- ◯ Mohale's Hoek
- ◯ Quthing
- ◯ Qacha's Nek
- ◯ Mokhotlong
- ◯ Thaba-Tseka

**Please enter the first name of your hair stylist**

______________________

**Please enter the last name (surname) of your hair stylist**

______________________

#### About yourself, more information

Remember, we are only interested in your information for research purposes and not to identify you personally. We will not share your personal information with anyone.

______________________

**What is the highest level of school/degree you have completed?**

- ◯ None
- ◯ Primary
- ◯ High School
- ◯ Vocational School
- ◯ Tertiary
- ◯ Prefer not to answer

**What is your relationship status?**

- ◯ Single
- ◯ Married
- ◯ Cohabitating
- ◯ Separated
- ◯ Divorced
- ◯ Widowed

**How many children do you have?**

- ◯ None
- ◯ 1
- ◯ 2
- ◯ 3
- ◯ 4
- ◯ More than 4

**Do you identify with any of the following religions?**

- ◯ Christianity
- ◯ Islam
- ◯ Other
- ◯ None
- ◯ Prefer not to answer

**Other, please describe**

______________________________________________

**Which of the following categories best describes your occupational status?**

- ◯ Employed/contractor
- ◯ Student
- ◯ Self-employed (own business)
- ◯ Housewife/Househusband
- ◯ No occupation at the moment
- ◯ Other
- ◯ Prefer not to answer

**Other, please describe**

______________________________________________

**How would you describe your financial situation these days?**

- ◯ Very comfortable
- ◯ Comfortable
- ◯ Just getting by
- ◯ Poor
- ◯ Very poor
- ◯ Prefer not to answer

#### Now, let's talk about your hair salon

**How often do you come to this hair salon?**

- ◯ More than twice a month
- ◯ Twice a month
- ◯ Once a month
- ◯ Once every two months
- ◯ Once every three months
- ◯ Less than once every three months

**How long does a typical visit at your hair salon last?**

- ◯ Less than an hour
- ◯ Between 1-2 hours
- ◯ Between 2-4 hours
- ◯ Between 4-6 hours
- ◯ More than 6 hours

**How long does it take you from home to get to your hair salon using your usual transport?**

- ◯ Less than 15min
- ◯ 15-30min
- ◯ 30-60min
- ◯ More than 1 hour

**How much money do you spend in transport (one way) to come to your hair salon using your usual transport?**

- ◯ M0-M10
- ◯ M10-20
- ◯ M20-30
- ◯ M30-more

**Are you (or your sexual partner/s) CURRENTLY using any contraception/family planning method?**

- ◯ Yes
- ◯ No
- ◯ Prefer not to answer

**Which method of contraception/family planning are you (or your sexual partner/s) currently using?**

*Select one or several answers*

- ☐ Male/external condoms
- ☐ Female/internal condoms
- ☐ Oral birth control pill
- ☐ Injectable (e.g., Depo-Provera)
- ☐ Implant (e.g., Implanon)
- ☐ Intrauterine device (IUD)
- ☐ Vaginal Ring
- ☐ Emergency contraception
- ☐ Rhythm (fertility awareness)
- ☐ Withdrawal
- ☐ Other

**Other, please describe your contraception/family planning method**

______________________________

**From where do you obtain your current contraception/family planning method?**

*Select one or several answers*

- ☐ Government health center/hospital
- ☐ Private health center/hospital
- ☐ Family planning clinic (e.g. LPPA)
- ☐ Pharmacy
- ☐ Community health worker/fieldworker
- ☐ Friend/relative
- ☐ Other

**Other, please describe where you obtain your current contraception/family planning method**

______________________________

#### Now, let's talk about your nearby health facility (health center or hospital)

**How long does it take you from home to go to your nearby hospital/health center using your usual transport?**

- ◯ Less than 15min
- ◯ 15-30min
- ◯ 30-60min
- ◯ More than 1 hour

**How much money do you spend on transport (one way) when you go to your nearby hospital/health center?**

- ◯ M0-M10
- ◯ M10-20
- ◯ M20-30
- ◯ M30-more

#### Now, let's talk about family planning methods

**How much do you agree with the following statement: If I wanted contraception/family planning, I know of a place where I can obtain it.**

- ◯ Strongly agree
- ◯ Agree
- ◯ Neither agree nor disagree
- ◯ Disagree
- ◯ Strongly disagree

**From where have you ever gotten information about contraception/family planning methods?**

*Select one or several answers*

- ☐ Health center/hospital
- ☐ From friends
- ☐ From family
- ☐ At school
- ☐ Internet/social media
- ☐ Never received any information
- ☐ Other place

**Other, please describe the location**

________________________________________

#### Now, let's talk about menstrual health products. These are products that help you to handle your period.

**Have you ever had a period/menstruation?**

- ◯ Yes
- ◯ No
- ◯ I prefer not to answer

##### » If you ever had a period/menstruation

**During your last menstrual period, what were all the materials you used to catch/absorb your menstruation when you were AT HOME?**

*Select one or several answers*

- ☐ Disposable sanitary pad
- ☐ Reusable (cloth) sanitary pad
- ☐ Tampon
- ☐ Cotton wool
- ☐ Toilet paper
- ☐ Cloth towel
- ☐ Natural material (e.g., leaves, sand, grass)
- ☐ Period underwear
- ☐ Underwear alone
- ☐ Mattress or foam
- ☐ Menstrual cup
- ☐ Other

**Other, please describe the product**

______________________________

**During your last menstrual period, what were all the materials you used to catch/absorb your menstruation when you were AWAY FROM HOME (e.g. at school/at work)?**

*Select one or several answers*

- [ ] Disposable sanitary pad
- [ ] Reusable (cloth) sanitary pad
- [ ] Tampon
- [ ] Cotton wool
- [ ] Toilet paper
- [ ] Cloth towel
- [ ] Natural material (e.g., leaves, sand, grass)
- [ ] Period underwear
- [ ] Underwear alone
- [ ] Mattress or foam
- [ ] Menstrual cup
- [ ] Other

**Other, please describe the product**

______________________________

**Would you like to learn more about reusable, washable, cloth sanitary pads for your period?**

- ○ Yes
- ○ No
- ○ I don't know what 'reusable, washable cloth sanitary pads' are

**Why not?**

*Select one or several answers*

- [ ] I think they are uncomfortable
- [ ] I think they are not hygienic
- [ ] I think they are not safe/absorbing enough
- [ ] I think they are expensive
- [ ] Other

**Other, please describe why not**

______________________________

**What do you think about the reusable, washable, cloth sanitary pads?**

*Select one or several answers*

- ☐ I think they are comfortable
- ☐ I think they are NOT comfortable
- ☐ I think they are hygienic
- ☐ I think they are NOT hygienic enough
- ☐ I think they are safe/absorbing enough
- ☐ I think they are NOT safe/absorbing enough
- ☐ I think they are cheap
- ☐ I think they are expensive
- ☐ Other

**Other, please describe**

______________________________

**Have you ever missed any school or work day due to your period/menstruation?**

- ○ Yes
- ○ No
- ○ I prefer not to answer

**How often in the past year?**

- ○ 1-2 times
- ○ 3-4 times
- ○ 5-6 times
- ○ More than 6 times

**What was the most common reason?**

- ○ Because of period pain/discomfort
- ○ Because I did not have access to menstruation health products
- ○ I prefer not to answer

**How old were you when you had your first period/menstruation?**

- ◯ Below 12 years of age
- ◯ Around 12 years of age
- ◯ Around 13 years of age
- ◯ Around 14 years of age
- ◯ Around 15 years of age
- ◯ Older than 15 years of age

#### Now, let's talk about HIV testing and Pre-Exposure Prophylaxis (PrEP) against HIV. PrEP is a pill you can take to prevent getting HIV.

**Have you ever heard of a pill, called pre-exposure prophylaxis (PrEP), to prevent getting HIV?**

- ◯ Yes
- ◯ No

How much do you agree with the following statements?

---

**PrEP is only for people who have many sexual partners**

- ◯ Strongly agree
- ◯ Agree
- ◯ Neither agree nor disagree
- ◯ Disagree
- ◯ Strongly disagree

**PrEP will cause people to have more risky sex**

- ◯ Strongly agree
- ◯ Agree
- ◯ Neither agree nor disagree
- ◯ Disagree
- ◯ Strongly disagree

**Only sex workers need PrEP**

- Strongly agree
- Agree
- Neither agree nor disagree
- Disagree
- Strongly disagree

**Only people with partner(s) living with HIV need PrEP**

- Strongly agree
- Agree
- Neither agree nor disagree
- Disagree
- Strongly disagree

**Instead of taking PrEP, people should just pick their partners carefully**

- Strongly agree
- Agree
- Neither agree nor disagree
- Disagree
- Strongly disagree

**Taking PrEP once provides lifelong protection against HIV**

- Strongly agree
- Agree
- Neither agree nor disagree
- Disagree
- Strongly disagree

**Taking PrEP prevents pregnancy**

- Strongly agree
- Agree
- Neither agree nor disagree
- Disagree
- Strongly disagree

**Taking PrEP protects me against other sexually transmitted infections such as syphilis**

- Strongly agree
- Agree
- Neither agree nor disagree
- Disagree
- Strongly disagree

**From where have you gotten information about PrEP?**

*Select one or several answers*

- Health center/hospital
- From friends
- From family
- At school
- Internet/social media
- Never received any information
- Other

**Other, please describe**

____________________________________________________

**How much do you agree with the following: I would be more likely to use PrEP if it also prevented pregnancy**

- Strongly agree
- Agree
- Neither agree nor disagree
- Disagree
- Strongly disagree

**Have you ever heard of the HIV self-test, a test that you can do yourself at home using a swab in your mouth to test for HIV?**

- Yes
- No

**What do you think your risk of getting HIV in the next year is?**

- ◯ No risk at all
- ◯ Small risk
- ◯ 50/50 (medium) risk
- ◯ High risk
- ◯ Very high risk
- ◯ I have been diagnosed with HIV
- ◯ I don't know
- ◯ I prefer not to answer

#### Now, let's talk about different Sexual and Reproductive Health (SRH) services. These are services to help you with your family planning needs, sexual well-being, and sexually transmitted diseases.

When answering the following questions, assume that you will receive all the services for free

---

How much do you agree with the following statements?

---

**I am comfortable receiving family planning/contraception counselling from a hair stylist in a hair salon.**

*This means receiving information to support young women in making decisions about having children or not, how many, and when, and where to obtain family planning methods.*

- ◯ Strongly agree
- ◯ Agree
- ◯ Neither agree nor disagree
- ◯ Disagree
- ◯ Strongly disagree

**Why do you disagree?**

*Select one or several answers*

- ☐ This is confidential, I don't want to talk about this in the hair salon with my hair stylist.
- ☐ Religious beliefs
- ☐ I don't think my hair stylist would be capable to provide this service
- ☐ I am not interested in such services at all
- ☐ I just prefer to receive this service from a healthcare professional
- ☐ Other reason

**Other reason, please describe**

---

**I am comfortable receiving male/external condoms (for free) from a hair stylist in a hair salon.**

*Male/external condoms are like protective shields that are worn on the penis during sex. They protect against pregnancy and sexually transmitted infections.*

- ◯ Strongly agree
- ◯ Agree
- ◯ Neither agree nor disagree
- ◯ Disagree
- ◯ Strongly disagree

**Why do you disagree?**

*Select one or several answers*

- ☐ This is confidential, I don't want to receive this in the hair salon
- ☐ Religious beliefs
- ☐ I don't think my hair stylist would be capable to provide this service
- ☐ I am not interested in such services at all
- ☐ I just prefer to receive this service from a healthcare professional
- ☐ Other reason

**Other reason, please describe**

---

**I am comfortable receiving female/internal condoms (for free) from a hair stylist in a hair salon.**

*Female/internal condoms are like protective shields that are inserted into the vagina during sex. They protect against pregnancy and sexually transmitted infections.*

- ◯ Strongly agree
- ◯ Agree
- ◯ Neither agree nor disagree
- ◯ Disagree
- ◯ Strongly disagree

**Why do you disagree?**

*Select one or several answers*

- [ ] This is confidential, I don't want to receive this in the hair salon
- [ ] Religious beliefs
- [ ] I don't think my hair stylist would be capable to provide this service
- [ ] I am not interested in such services at all
- [ ] I just prefer to receive this service from a healthcare professional
- [ ] Other reason

**Other reason, please describe**

---

**I am comfortable receiving the oral birth control pill (for free) from a hair stylist in a hair salon.**

*Oral birth control pills are a type of medicine that women take daily by mouth to prevent pregnancy.*

- ◯ Strongly agree
- ◯ Agree
- ◯ Neither agree nor disagree
- ◯ Disagree
- ◯ Strongly disagree

**Why do you disagree?**

*Select one or several answers*

- [ ] This is confidential, I don't want to receive this in the hair salon
- [ ] Religious beliefs
- [ ] I don't think my hair stylist would be capable to provide this service
- [ ] I am not interested in such services at all
- [ ] I just prefer to receive this service from a healthcare professional
- [ ] Other reason

**Other reason, please describe**

---

**I am comfortable receiving the emergency contraception pill (for free) from a hair stylist in a hair salon.**

*Emergency contraception pills are a type of medicine that women can take right after unprotected sex to prevent a pregnancy.*

- ◯ Strongly agree
- ◯ Agree
- ◯ Neither agree nor disagree
- ◯ Disagree
- ◯ Strongly disagree

**Why do you disagree?**

*Select one or several answers*

- [ ] This is confidential, I don't want to receive this in the hair salon
- [ ] Religious beliefs
- [ ] I don't think my hair stylist would be capable to provide this service
- [ ] I am not interested in such services at all
- [ ] I just prefer to receive this service from a healthcare professional
- [ ] Other reason

**Other reason, please describe**

______________________________

**I am comfortable receiving HIV counselling from a hair stylist in a hair salon.**

*This means receiving information about how HIV can spread and steps that can be taken to protect oneself and others, as well as information about where to obtain more information and testing possibilities.*

- ○ Strongly agree
- ○ Agree
- ○ Neither agree nor disagree
- ○ Disagree
- ○ Strongly disagree

**Why do you disagree?**

*Select one or several answers*

- [ ] This is confidential, I don't want to talk about this in the hair salon with my hair stylist.
- [ ] Religious beliefs
- [ ] I don't think my hair stylist would be capable to provide this service
- [ ] I am not interested in such services at all
- [ ] I just prefer to receive this service from a healthcare professional
- [ ] Other reason

**Other reason, please describe**

______________________________

**I am comfortable receiving a test kit for oral HIV self-testing (for free) from a hair stylist in a hair salon.**

*A test kit for oral HIV self-testing includes a swab that allows you to collect fluid yourself from inside your mouth to test for HIV. It is designed to allow you to take the HIV test in private and anonymously.*

- ◯ Strongly agree
- ◯ Agree
- ◯ Neither agree nor disagree
- ◯ Disagree
- ◯ Strongly disagree

**Why do you disagree?**

*Select one or several answers*

- ☐ This is confidential, I don't want to receive this in the hair salon
- ☐ Religious beliefs
- ☐ I don't think my hair stylist would be capable to provide this service
- ☐ I am not interested in such services at all
- ☐ I just prefer to receive this service from a healthcare professional
- ☐ Other reason

**Other reason, please describe**

______________________________

**I am comfortable receiving pre-exposure prophylaxis (PrEP), for free, from a hair stylist in a hair salon.**

*PrEP is a pill that you can take to prevent getting HIV if you are exposed to it.*

- ◯ Strongly agree
- ◯ Agree
- ◯ Neither agree nor disagree
- ◯ Disagree
- ◯ Strongly disagree

**Why do you disagree?**

*Select one or several answers*

- ☐ This is confidential, I don't want to receive this in the hair salon
- ☐ Religious beliefs
- ☐ I don't think my hair stylist would be capable to provide this service
- ☐ I am not interested in such services at all
- ☐ I just prefer to receive this service from a healthcare professional
- ☐ Other reason

**Other reason, please describe**

______________________________

**I am comfortable receiving post-exposure prophylaxis (PEP), for free, from a hair stylist in a hair salon.**

*PEP is a pill you can take if you think you have been exposed to HIV. It's like an emergency treatment that helps prevent the virus from spreading in your body.*

- ◯ Strongly agree
- ◯ Agree
- ◯ Neither agree nor disagree
- ◯ Disagree
- ◯ Strongly disagree

**Why do you disagree?**

*Select one or several answers*

- ☐ This is confidential, I don't want to receive this in the hair salon
- ☐ Religious beliefs
- ☐ I don't think my hair stylist would be capable to provide this service
- ☐ I am not interested in such services at all
- ☐ I just prefer to receive this service from a healthcare professional
- ☐ Other reason

**Other reason, please describe**

______________________________________________

**I am comfortable receiving counselling about Sexually Transmitted Infections (STIs/STDs) from a hair stylist in a hair salon.**

*This means receiving information about how STIs can spread and steps that can be taken to protect yourself and your partner. And information about where to obtain more information and testing possibilities.*

- ◯ Strongly agree
- ◯ Agree
- ◯ Neither agree nor disagree
- ◯ Disagree
- ◯ Strongly disagree

**Why do you disagree?**

*Select one or several answers*

- ☐ This is confidential, I don't want to talk about this in the hair salon with my hair stylist.
- ☐ Religious beliefs
- ☐ I don't think my hair stylist would be capable to provide this service
- ☐ I am not interested in such services at all
- ☐ I just prefer to receive this service from a healthcare professional
- ☐ Other reason

**Other reason, please describe**

______________________________

**I am comfortable receiving counselling about menstrual health from a hair stylist in a hair salon.**

*This means receiving information about how to handle your period and hygiene around menstrual health. And information about where to obtain menstrual products.*

- ◯ Strongly agree
- ◯ Agree
- ◯ Neither agree nor disagree
- ◯ Disagree
- ◯ Strongly disagree

**Why do you disagree?**

*Select one or several answers*

- ☐ This is confidential, I don't want to talk about this in the hair salon with my hair stylist.
- ☐ Religious beliefs
- ☐ I don't think my hair stylist would be capable to provide this service
- ☐ I am not interested in such services at all
- ☐ I just prefer to receive this service from a healthcare professional
- ☐ Other reason

**Other reason, please describe**

______________________________

**I am comfortable receiving menstrual products (for example reusable cloth sanitary pads), for free, from a hair stylist in a hair salon.**

*Menstrual products such as the reusable cloth sanitary pads absorb the menstrual flow and help you to stay clean and comfortable during your period.*

- ◯ Strongly agree
- ◯ Agree
- ◯ Neither agree nor disagree
- ◯ Disagree
- ◯ Strongly disagree

**Why do you disagree?**

*Select one or several answers*

- [ ] This is confidential, I don't want to receive this in the hair salon
- [ ] Religious beliefs
- [ ] I don't think my hair stylist would be capable to provide this service
- [ ] I am not interested in such services at all
- [ ] I just prefer to receive this service from a healthcare professional
- [ ] Other reason

**Other reason, please describe**

---

**I am comfortable receiving gender-based violence (GBV) information from a hair stylist in a hair salon.**

*This means having a space for people who have experienced violence or abuse based on their gender to talk about it. And information about where to get further help.*

- ○ Strongly agree
- ○ Agree
- ○ Neither agree nor disagree
- ○ Disagree
- ○ Strongly disagree

**Why do you disagree?**

*Select one or several answers*

- [ ] This is confidential, I don't want to talk about this in the hair salon with my hair stylist.
- [ ] Religious beliefs
- [ ] I don't think my hair stylist would be capable to provide this service
- [ ] I am not interested in such services at all
- [ ] I just prefer to receive this service from a healthcare professional
- [ ] Other reason

**Other reason, please describe**

---

**Which of the following Sexual and Reproductive Health (SRH) services would you be interested in receiving *if your hair stylist provided it in the hair salon in the next 6 months?***

*Select one or several answers*

- [ ] Family planning/contraception counselling
- [ ] Male/external condoms
- [ ] Female/internal condoms
- [ ] Oral birth control pill
- [ ] Emergency contraception pill
- [ ] HIV counselling
- [ ] HIV testing using oral HIV self-testing kits
- [ ] Pre-exposure prophylaxis (PrEP)
- [ ] Post-exposure prophylaxis (PEP)
- [ ] Sexually Transmitted Infections (STIs/STDs) counselling
- [ ] Menstrual health counselling
- [ ] Menstrual products (for example reusable cloth sanitary pads)
- [ ] Gender-based violence information
- [ ] None of them

#### Thank you very much

Once you submit this survey, we will check your answers and send you a small gift to your Mpesa. This may take a few hours/days.

---

If you have any questions, please call the HUB (+266 5708 7179).

---

Please sign on the next screen and submit the survey.

---

**Please sign here. Thank you very much for your participation.**

*Draw your signature with your finger*

Text S6. Interview guides.

**For hair stylists:**

| SECTION ONE : STYLIST AS SCIENTIST | |
|---|---|
| ***This section aims to hear about your experience with the survey and being involved in this research*** | |
| ***Questions*** | ***Probes*** |
| How did you feel about your involvement (recruiting clients & assisting with survey) in this research project? | ▪ Were you comfortable telling clients about the survey?<br>▪ Are there parts of this process that were confusing?<br>▪ Are there parts of it that you found easy/intuitive?<br>▪ If we could do things differently, what would you like to change? |
| What did you think about the reimbursement? | ▪ Did you think the reimbursement was fair?<br>▪ What about the payment process itself? Using MPesa, timeline, |
| **SECTION TWO: UNDERSTANDING OF SRH AND ITS COMPONENTS** | |
| ***This section intends to broadly introduce the topics. The assumption is that many of the components will be familiar from the survey process*** | |
| ***Questions*** | ***Probes*** |
| What comes to mind when we mention sexual and reproductive health services? | *Follow up questions and probes*<br>▪ Have you used/needed these services before? If yes, where did you find them? |
| Have you ever provided/sold menstrual health products or condoms in your salon? | *Follow up questions and probes*<br>▪ If yes, in what instances did you provide the products?/ Do you still sell the products?<br>▪ If no, is this something you have considered? |
| **SECTION THREE: CONTRACEPTIVES, PEP, GBV** | |
| ***This section may include sensitive topics*** | |
| Now, I would like to speak about family planning and contraceptive services | |

| ***Questions*** | ***Probes*** |
|---|---|
| How would you feel about providing family planning counseling and condoms to your clients? | *Follow up questions and probes*<br>▪ What would be your concerns?/ What aspects would you not be comfortable with?<br>▪ How would you want to be supported?/ What help would you need? |
| How would you feel about providing the oral birth control pill? | *Follow up questions and probes*<br>▪ What would be your concerns?/ What aspects would you not be comfortable with?<br>▪ How would you want to be supported?/ What help would you need? |
| How would you feel about providing the **emergency** birth control pill? (plan B) | *Follow up questions and probes*<br>▪ What would be your concerns?/ What aspects would you not be comfortable with?<br>▪ How would you want to be supported?/ What help would you need? |
| How would you feel about providing Post Exposure prophylaxis – or PEP? | *Follow up questions and probes*<br>▪ What would be your concerns?<br>▪ What can be done to support you if you were to provide PEP? |
| Would you be comfortable speaking to your clients about an incidence of gender-based violence they experienced/witnessed? | Follow up questions and probes<br>▪ What would make it easy to speak to your clients about GBV?<br>▪ What would be your concerns?/ What aspects would you not be comfortable with? |

**SECTION FOUR: HIVST & PREVENTION SERVICES**

***This section is intended to hear about comfort with providing information about HIV preventative services***

*Add transitional statement: Thank you for sharing your thoughts. Now I would like us to talk about PrEP and PEP, which are both ways of preventing a person from acquiring HIV*

| ***Questions*** | ***Probes*** |
|---|---|
| Are you familiar with HIV self-testing? | *Follow up questions and probes*<br>▪ If yes, how did you learn about it?<br>▪ *If no, explain how it works*<br>▪ Would you be comfortable offering the oral self-test in your salon? |

| | |
|---|---|
| | ▪ What would be your concerns?/ What aspects would you not be comfortable with?<br>▪ How would you want to be supported?/ What help would you need? |
| Have you heard about PrEP? | *Follow up questions and probes*<br>▪ Where have you heard about PrEP?<br>▪ What is your understanding of how it works?<br>▪ How would you feel about providing PrEP information and supplies to clients for free? |
| **SECTION FIVE: MENSTRUAL HEALTH** | |
| ***This section is intended to hear about comfort with providing information about and access to menstrual health products to your client***<br>Next, I want us to talk about menstrual health | |
| ***Questions*** | ***Probes*** |
| How would you feel about providing menstrual health products for free to your clients? | *Follow up questions and probes*<br>▪ What would be the advantage of providing these products?<br>▪ What would be your concerns?/ What aspects would you not be comfortable with? |
| If you received training, how would you feel about providing information about, and supplies of the reusable pad to your clients? | *Follow up questions and probes*<br>▪ What would be your concerns?/ What aspects would you not be comfortable with?<br>▪ How would you want to be supported?/ What help would you need? |

**Thank the participants for their time**

**<u>For clients:</u>**

| SECTION ONE: UNDERSTANDING OF SRH AND ITS COMPONENTS | |
|---|---|
| ***This section intends to establish rapport with the declarants, and to establish their understanding about sexual and reproductive health*** | |
| *Transitional/introductory statement for flow:* | |
| ***<u>Questions</u>*** | ***<u>Probes</u>*** |
| What comes to mind when we mention sexual and reproductive health services? | *Follow up questions and probes*<br>▪ Have you used/needed these services before? If yes, where did you find them?<br>▪ If someone you knew needed said services, where would you advise them to go? |
| Have you faced any challenges accessing these services when you needed them? | *Follow up questions and probes*<br>▪ Would you mind sharing what challenges you faced?<br>▪ If not, what made it easy to get these services?<br>▪ Are there any people you relied on to help you with accessing these services? Why them? |
| **SECTION TWO: CONTRACEPTIVES/FAMILY PLANNING AND PEP** | |
| ***After section one, introduce the hair salon project and its aims*** | |
| Now, I would like to speak about family planning and contraceptive services | |
| ***<u>Questions</u>*** | ***<u>Probes</u>*** |
| How would you feel about getting family planning services (for example, male/female condom) from your hair stylist? | *Follow up questions and probes*<br>▪ What would be the advantage of getting these services from your stylist?<br>▪ What challenges would you expect?/ What aspects would you not be comfortable with? |
| Are you familiar with the oral birth control pill and how it works? | *Follow up questions and probes*<br>▪ *If yes, keep going. If not, explain how it works.*<br>▪ What would be the advantage of getting the pill from your stylist?<br>▪ What challenges would you expect?/ What aspects would you not be comfortable with? |

| | |
|---|---|
| Are you familiar with the emergency pill (plan B) and how it works? | *Follow up questions and probes*<br>▪ *If yes, keep going. If not, explain how it works.*<br>▪ What would be the advantage of getting the pill from your stylist?<br>▪ What challenges would you expect?/ What aspects would you not be comfortable with? |
| Are you familiar with PEP – Post Exposure Prophylaxis and how it works? | *Follow up questions and probes*<br>▪ *If yes, keep going. If not, explain how it works.*<br>▪ What would be the advantage of getting PEP from your stylist?<br>▪ What challenges would you expect?/ What aspects would you not be comfortable with? |

#### SECTION THREE: HIVST & PREP

***This section is intended to hear about comfort with getting information about HIV preventative services***

*Add transitional statement: Thank you for sharing your thoughts. Now I would like us to talk about PrEP and PEP, which are both ways of preventing a person from acquiring HIV*

| ***Questions*** | ***Probes*** |
|---|---|
| Are you familiar with HIV self-testing? | *Follow up questions and probes*<br>▪ If yes, how did you learn about it?<br>▪ *If no, explain how it works*<br>▪ Would you be comfortable getting the oral self-test from your salon?<br>▪ What challenges would you expect?/ What aspects would you not be comfortable with? |
| Have you heard about PrEP? | *Follow up questions and probes*<br>▪ Where have you heard about it?<br>▪ What is your understanding of how it works?<br>▪ Do you know where to find PrEP when you need it?<br>*Only if this is available in Lesotho at the time:*<br>▪ Would you be interested in getting the injectable PrEP? |
| How would you feel about getting PrEP from your hair stylist? | *Follow up questions and probes*<br>▪ What would you appreciate about getting (regular pill) PrEP from your stylist? |

| | |
|---|---|
| | ▪ What would make you uncomfortable?<br>▪ What can your stylist do to make you feel comfortable enough to ask questions about PrEP? |
| **SECTION FOUR: MENSTRUAL HEALTH** | |
| ***This section is intended to hear about comfort with getting information about and access to menstrual health products from your hair stylist***<br>Next, I want us to talk about menstrual health | |
| ***<u>Questions</u>*** | ***<u>Probes</u>*** |
| How would you feel about getting menstrual health products for free from your hair stylist? | *Follow up questions and probes*<br>▪ What would be the advantage of getting these products from your stylist?<br>▪ What challenges would you expect?/ What aspects would you not be comfortable with? |
| Are you familiar with the reusable menstrual pad? | *Follow up questions and probes*<br>▪ Where have you heard about it?<br>▪ What is your understanding of how it works?<br>▪ Would you be comfortable using the reusable pad? Why or why not?<br>▪ Would you be comfortable getting the reusable pads from your stylist? Why or why not? |

| | |
|---|---|
| **SECTION FIVE: GENDER-BASED VIOLENCE** | |
| ***This section is intended to hear about comfort with getting information about and access to gender-based violence support from one's stylist*** | |
| ***<u>Questions</u>*** | ***<u>Probes</u>*** |
| What is your understanding of gender-based violence? | Follow up questions and probes<br>▪ What does it include?<br>▪ Do you feel like you know where to get resources about GBV? |
| Where would you ideally want to get resources or information about GBV? | Follow up questions and probes<br>▪ Which people do you think you would feel comfortable reaching out to? Why them?<br>▪ What support would you want to receive? From whom?<br>▪ What can your stylist do to make you feel comfortable enough to ask questions about gender based violence? |

**Thank the participants for their time**

#### Figure S1. Overview of survey steps and procedures

The HUB distributes the survey recruitment webpage (www.hairsalonproject.com) via social media.

↓

Stylist accesses webpage on smartphone, reads info material and confirms eligibility criteria.

↓

Stylist registers themself and their hair salon via registration questionnaire asking the following:

1) Address & name of hair salon
2) GPS coordinates
3) Picture of hair salon (outside and inside)
4) WhatsApp phone number
5) Mpesa phone number

→ The HUB verifies all information in real-time: Is the hair salon located in Lesotho? Duplicate? Does the Mpesa account correspond to the name? All eligibility criteria fulfilled?

↓

Verified and eligible stylist receives the consent form and the stylist questionnaire (KoboCollect link) through WhatsApp, provides consent, fills in the questionnaire and uploads all

→ The HUB confirms the receipt of the questionnaire and asks the stylist to recruit up to three clients and, for this, sends the link of the client registration questionnaire to the stylist via WhatsApp.

↓

Stylist recruits a client who uses the link (on the stylists' or clients' phone), to fill in the client registration questionnaire (e.g. during a hair salon session).

↓

Client registers herself via registration questionnaire asking the following:

1) Address & name of hair salon
2) GPS coordinates
3) "Selfie at the hair salon" picture
4) WhatsApp phone number
5) Mpesa phone number

→ The HUB verifies all information in real-time: Is the hair salon located in Lesotho? Duplicate? Does the Mpesa account correspond to the name? All eligibility criteria fulfilled?

↓

Verified and eligible client receives the consent form and the client questionnaire (KoboCollect link) through WhatsApp, provides consent, fills in the questionnaire and uploads all.

→ The HUB confirms the receipt of the questionnaire and links the client to the respective stylist.

↓

Client receives token of appreciation via Mpesa.

Stylist receives remuneration for work via Mpesa and a Citizen Scientist certificate.

The stylist received 100 LSL (5.8 USD) via M-PESA per successfully submitted client questionnaire to reimburse their time and they received a citizen scientist certificate for participation. The clients received a token of appreciation of the value of a basic hair style via M-PESA upon completion of the questionnaire (cornrows; 50 LSL, ca. 2.9 USD).

Figure S2. Participants flowchart.

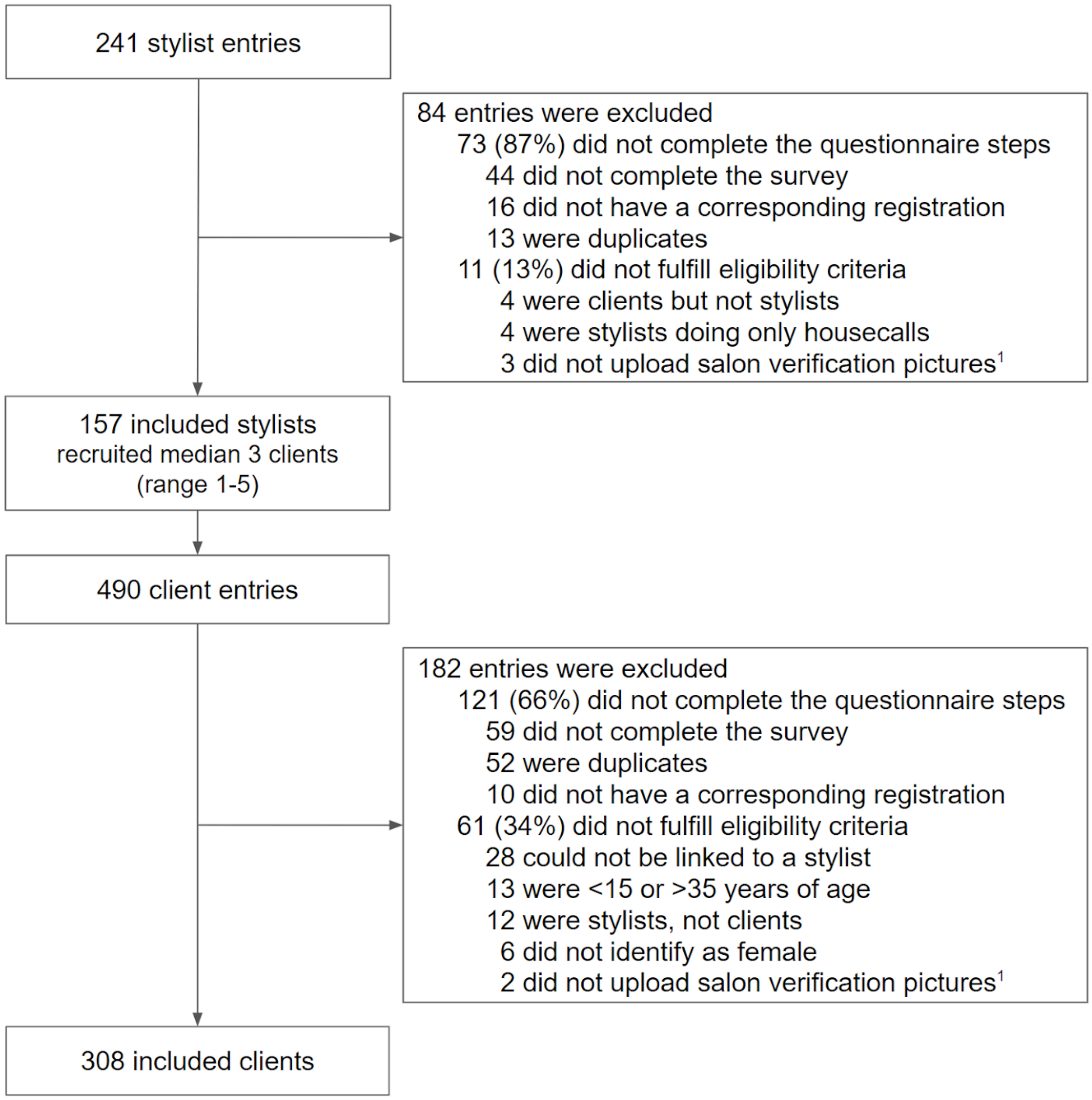


[1] Despite multiple contact attempts, no photos were uploaded, preventing the research team from verifying the existence of a hair salon

Figure S3. Map of the participating 157 hair salons/stylists

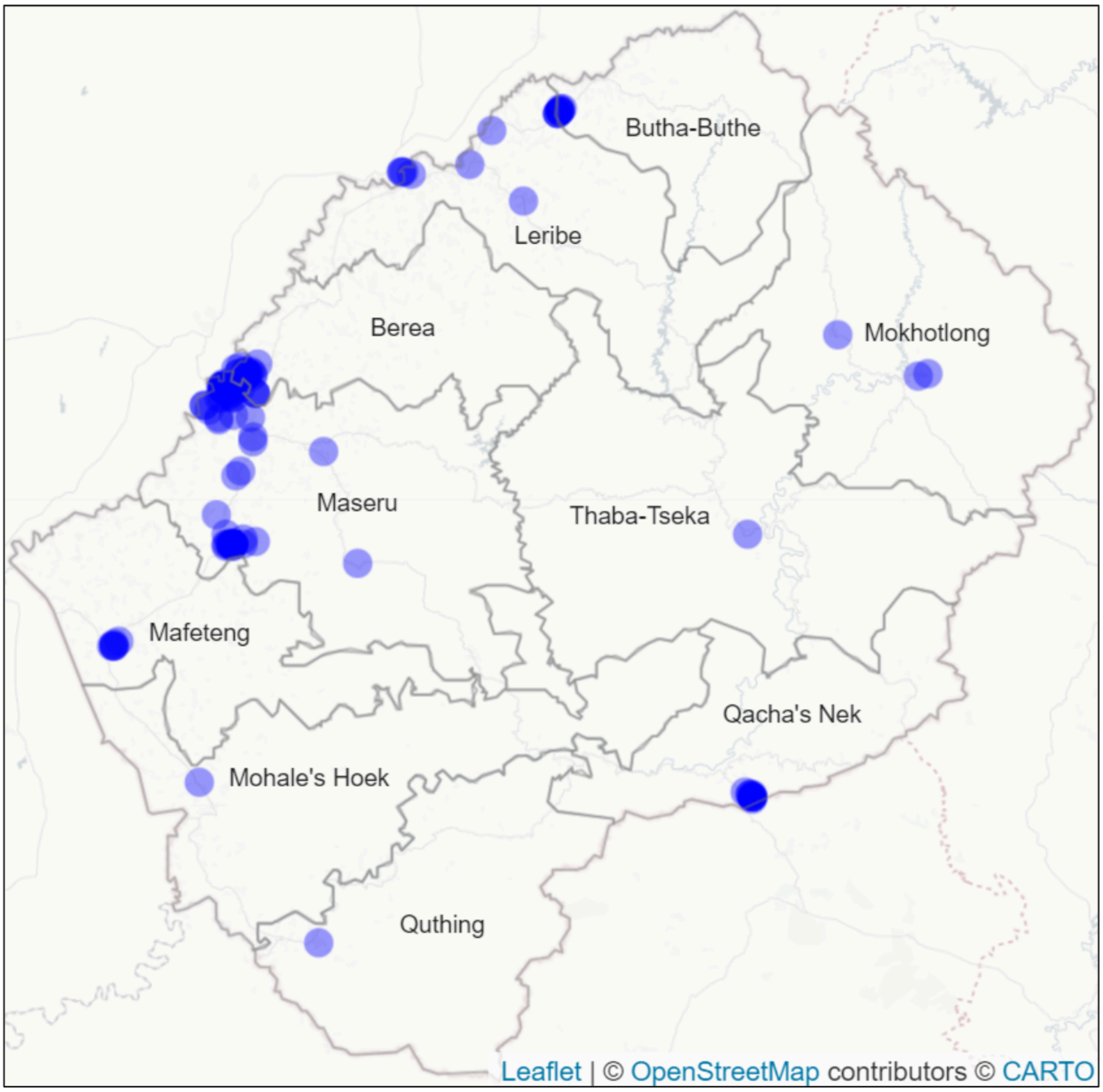


We recruited hair salons/stylists across all 10 districts of Lesotho: Berea 8 (5.1%), Butha-Buthe 8 (5.1%), Leribe 9 (5.7%), Mafeteng 8 (5.1%), Maseru 107 (68.2%), Mohales Hoek 1 (0.6%), Mokhotlong 4 (2.5%), Qacha's Nek 10 (6.4%), Quthing 1 (0.6%) and Thaba-Tseka 1 (0.6%).

#### Table S1. Interviewed stylists and clients' baseline characteristics.

| | | **Stylists** | **Clients** |
|---|---|---|---|
| | | **n = 6** | **n = 8** |
| Gender (%) | Woman/girl | 5 ( 83.3) | 8 (100) |
| | Man/boy | 1 ( 16.7) | |
| Age (median [IQR]) | | 26.50 [26.00, 28.50] | 22.50 [20.75, 25.75] |
| Education (%) | Primary | 1 ( 16.7) | 0 ( 0.0) |
| | High school | 1 ( 16.7) | 3 ( 37.5) |
| | Vocational school | 1 ( 16.7) | 2 ( 25.0) |
| | Tertiary | 3 ( 50.0) | 2 ( 25.0) |
| | Prefer not to answer | 0 ( 0.0) | 1 ( 12.5) |
| Religion (%) | Christianity | 5 ( 83.3) | 6 ( 75.0) |
| | Prefer not to answer | 1 ( 16.7) | 2 ( 25.0) |
| Owner of the hair salon (%) | Yes | 5 ( 83.3) | |
| | No | 1 ( 16.7) | |
| Work experience as a stylist (%) | Up to 1 year | 1 ( 16.7) | |
| | 1 to 3 years | 2 ( 33.3) | |
| | 3 to 6 years | 0 ( 0.0) | |
| | More than 6 years | 3 ( 50.0) | |
| Relationship status (%) | Single | | 6 ( 75.0) |
| | Married | | 1 ( 12.5) |
| | Cohabiting | | 1 ( 12.5) |
| Number of children (%) | None | | 7 ( 87.5) |
| | 1 | | 0 ( 0.0) |
| | 2 | | 0 ( 0.0) |
| | 3 | | 1 ( 12.5) |
| Economical status (%) | Very poor | | 1 ( 12.5) |
| | Poor | | 0 ( 0.0) |
| | Getting by | | 4 ( 50.0) |
| | Comfortable | | 2 ( 25.0) |
| | Prefer not to answer | | 1 ( 12.5) |
| Occupation (%) | Employed | | 3 ( 37.5) |
| | Self-employed | | 1 ( 12.5) |
| | Homemaker | | 1 ( 12.5) |
| | Student | | 2 ( 25.0) |
| | None | | 1 ( 12.5) |

| | | | |
|---|---|---|---|
| Comfortable offering/receiving PrEP or similar services (%) | Yes | 4 (66.7) | 6 (75.0) |
| | No | 2 (33.3) | 2 (25.0) |

#### Table S2. PrEP, HIVST, Family planning and Menstrual health.

| | | Stylists | Clients |
|---|---|---|---|
| Ever heard about HIV self-test[1] (%) | Yes | 153 (97.5) | 297 (96.4) |
| | No | 4 ( 2.5) | 11 ( 3.6) |
| Ever heard about PrEP[1] (%) | Yes | 144 (91.7) | 255 (82.8) |
| | No | 13 ( 8.3) | 53 (17.2) |
| Agreement with the following myths about PrEP[2] (%) | PrEP is only for people who have many sexual partners | 44 (30.6) | 52 (20.4) |
| | PrEP will cause people to have more risky sex | 59 (41.0) | 109 (42.7) |
| | Only sex workers need PrEP | 16 (11.1) | 13 ( 5.1) |
| | Only people with partner(s) living with HIV need PrEP | 27 (18.8) | 31 (12.2) |
| | Instead of taking PrEP, people should just pick their partners carefully | 38 (26.4) | 57 (22.4) |
| | Taking PrEP once provides lifelong protection against HIV | 44 (30.6) | 90 (35.3) |
| | Taking PrEP prevents pregnancy | 4 ( 2.8) | 4 ( 1.6) |
| | Taking PrEP protects me against other sexually transmitted infections such as syphilis | 22 (15.3) | 34 (13.3) |
| Currently used contraceptive method[3] (%) | External condom | 32 ( 32.3) | 76 ( 45.0) |
| | Internal condom | 5 ( 5.1) | 12 ( 7.1) |
| | Oral birth control pill | 31 ( 31.3) | 42 ( 24.9) |
| | Injectable | 23 ( 23.2) | 42 ( 24.9) |
| | Implant | 7 ( 7.1) | 8 ( 4.7) |
| | Intrauterine device | 3 ( 3.0) | 6 ( 3.6) |
| | Vaginal ring | 0 (0.0) | 0 (0.0) |
| | Emergency contraception pill | 1 ( 1.0) | 1 ( 0.6) |
| | Rhythm | 1 ( 1.0) | 1 ( 0.6) |
| | Withdrawal | 2 ( 2.0) | 6 ( 3.6) |
| Source of current contraception method[3] (%) | Public health center | 48 (48.5) | 90 (53.3) |
| | Private health center | 8 ( 8.1) | 7 ( 4.1) |
| | Family planning clinic | 6 ( 6.1) | 17 (10.1) |
| | Pharmacy | 38 (38.4) | 62 (36.7) |
| | Community helath worker | 3 ( 3.0) | 2 ( 1.2) |
| | Friend/relative | 2 ( 2.0) | 1 ( 0.6) |
| | Other | 2 ( 2.0) | 4 ( 2.4) |
| Materials used during last menstrual period[4] (%) | Disposable sanitary pad | 134 ( 91.8) | 270 ( 92.5) |
| | Tampon | 12 ( 8.2) | 22 ( 7.5) |

| | | | |
|---|---|---|---|
| | Toilet paper | 5 ( 3.4) | 6 ( 2.1) |
| | Reusable (cloth) sanitary pad | 2 ( 1.4) | 9 ( 3.1) |
| | Cotton wool | 0 ( 0.0) | 3 ( 1.0) |
| | Cloth towel | 2 ( 1.4) | 2 ( 0.7) |
| | Period underwear | 2 ( 1.4) | 1 ( 0.3) |
| | Underwear alone | 1 ( 0.7) | 1 ( 0.3) |
| | Menstrual cup | 1 ( 0.7) | 1 ( 0.3) |
| Menarche age[4] (%) | Below 12 years of age | 10 ( 6.8) | 25 ( 8.6) |
| | Around 12 years of age | 25 (17.1) | 64 (21.9) |
| | Around 13 years of age | 37 (25.3) | 56 (19.2) |
| | Around 14 years of age | 29 (19.9) | 65 (22.3) |
| | Around 15 years of age | 25 (17.1) | 52 (17.8) |
| | Older than 15 years of age | 20 (13.7) | 30 (10.3) |
| Missed at least one school or work day due to menstruation[4] (%) | Yes | 43 (29.5) | 72 (24.7) |
| | No | 101 (69.2) | 219 (75.0) |
| | Prefer not to answer | 2 ( 1.4) | 2 ( 1.4) |
| Frequency of days missed during the last year due to menstruation[5] (%) | 1-2 times | 19 (44.2) | 36 (50.0) |
| | 3-4 times | 15 (34.9) | 21 (29.2) |
| | 5-6 times | 1 ( 2.3) | 3 ( 4.2) |
| | More than 6 times | 8 (18.6) | 12 (16.7) |
| Reasons for days missed during the last year due to menstruation[5] (%) | No access to menstruation health products | 6 (14.0) | 10 (13.9) |
| | Period pain/discomfort | 33 (76.7) | 61 (84.7) |
| | Prefer not to answer | 4 ( 9.3) | 1 ( 1.4) |

[1] Assessed among all stylists and clients (Stylists: n = 157, Clients: n = 308)

[2] Assessed only among those that have heard about PrEP (Stylists: n = 144, Clients: n = 255)

[3] Only among those currently using a contraceptive method (Stylists: n = 99, Clients: n = 169)

[4] Only among those who experienced a menstrual period (Stylists: n = 146, Clients: n = 292)

[5] Only among those who ever missed at least any school or work day due to menstruation (Stylists: n = 43, Clients: n = 72)

Table S3. Comfortability of stylists offering sexual and reproductive health services.

| n = 157 | Comfortable | Neutral position | Not comfortable |
|---|---|---|---|
| Family planning counselling (%) | 143 (91.1) | 6 (3.8) | 8 (5.1) |
| External/male condoms (%) | 148 (94.3) | 3 (1.9) | 6 (3.8) |
| Internal condoms/female condoms (%) | 148 (94.3) | 3 (1.9) | 6 (3.8) |
| Contraceptive pill (%) | 146 (93) | 4 (2.5) | 7 (4.5) |
| Emergency contraceptive pill (%) | 145 (92.4) | 6 (3.8) | 6 (3.8) |
| HIV counselling (%) | 136 (86.6) | 9 (5.7) | 12 (7.6) |
| Oral HIV self-testing (%) | 147 (93.6) | 5 (3.2) | 5 (3.2) |
| HIV pre-exposure prophylaxis (%) | 145 (92.4) | 5 (3.2) | 7 (4.5) |
| HIV post-exposure prophylaxis (%) | 144 (91.7) | 7 (4.5) | 6 (3.8) |
| Sexually transmitted infection counselling (%) | 137 (87.3) | 8 (5.1) | 12 (7.6) |
| Menstrual counselling (%) | 148 (94.3) | 4 (2.5) | 5 (3.2) |
| Menstrual products (%) | 151 (96.2) | 3 (1.9) | 3 (1.9) |
| Gender-based violence information (%) | 149 (94.9) | 3 (1.9) | 5 (3.2) |

Table S4. Comfortability of clients receiving sexual and reproductive health services.

| | | Overall | | | 15-24 yrs | | | 25-35 yrs | |
|---|---|---|---|---|---|---|---|---|---|
| | | n = 308 | | | n = 128 | | | n = 180 | |
| | Comfortable | Neutral position | Not comfortable | Comfortable | Neutral position | Not comfortable | Comfortable | Neutral position | Not comfortable |
| Family planning counselling (%) | 277 (89.9) | 7 (2.3) | 24 (7.8) | 116 (90.6%) | 3 (2.3%) | 9 (7%) | 161 (89.4%) | 4 (2.2%) | 15 (8.3%) |
| External/male condoms (%) | 271 (88) | 17 (5.5) | 20 (6.5) | 103 (80.5%) | 13 (10.2%) | 12 (9.4%) | 168 (93.3%) | 4 (2.2%) | 8 (4.4%) |
| Internal condoms/female condoms (%) | 277 (89.9) | 10 (3.2) | 21 (6.8) | 110 (85.9%) | 7 (5.5%) | 11 (8.6%) | 167 (92.8%) | 3 (1.7%) | 10 (5.6%) |
| Contraceptive pill (%) | 276 (89.6) | 5 (1.6) | 27 (8.8) | 113 (88.3%) | 2 (1.6%) | 13 (10.2%) | 163 (90.6%) | 3 (1.7%) | 14 (7.8%) |
| Emergency contraceptive pill (%) | 279 (90.6) | 9 (2.9) | 20 (6.5) | 113 (88.3%) | 6 (4.7%) | 9 (7%) | 166 (92.2%) | 3 (1.7%) | 11 (6.1%) |
| HIV counselling (%) | 257 (83.4) | 15 (4.9) | 36 (11.7) | 102 (79.7%) | 9 (7%) | 17 (13.3%) | 155 (86.1%) | 6 (3.3%) | 19 (10.6%) |
| Oral HIV self-testing (%) | 288 (93.5) | 6 (1.9) | 14 (4.5) | 118 (92.2%) | 3 (2.3%) | 7 (5.5%) | 170 (94.4%) | 3 (1.7%) | 7 (3.9%) |
| HIV pre-exposure prophylaxis (%) | 272 (88.3) | 16 (5.2) | 20 (6.5) | 111 (86.7%) | 9 (7%) | 8 (6.2%) | 161 (89.4%) | 7 (3.9%) | 12 (6.7%) |
| HIV post-exposure prophylaxis (%) | 266 (86.4) | 16 (5.2) | 26 (8.4) | 108 (84.4%) | 9 (7%) | 11 (8.6%) | 158 (87.8%) | 7 (3.9%) | 15 (8.3%) |
| Sexually transmitted infection counselling (%) | 262 (85.1) | 12 (3.9) | 34 (11) | 107 (83.6%) | 3 (2.3%) | 18 (14.1%) | 155 (86.1%) | 9 (5%) | 16 (8.9%) |
| Menstrual counselling (%) | 281 (91.2) | 8 (2.6) | 19 (6.2) | 118 (92.2%) | 2 (1.6%) | 8 (6.2%) | 163 (90.6%) | 6 (3.3%) | 11 (6.1%) |
| Menstrual products (%) | 294 (95.5) | 5 (1.6) | 9 (2.9) | 121 (94.5%) | 3 (2.3%) | 4 (3.1%) | 173 (96.1%) | 2 (1.1%) | 5 (2.8%) |
| Gender-based violence information (%) | 287 (93.2) | 6 (1.9) | 15 (4.9) | 117 (91.4%) | 2 (1.6%) | 9 (7%) | 170 (94.4%) | 4 (2.2%) | 6 (3.3%) |

Table S5. Barriers and facilitators of offering/receiving HIV/SRH services at hair salons. Qualitative analysis matrix.

| Barriers - Stylists | | |
|---|---|---|
| **Themes** | **Codes** | **Illustrative quotes** |
| **Problems with current infrastructure** | Concerns about safe storage | "You know how sometimes pills are kept in a big container for safe storage, so the hair stylists may not own the needed storage equipment, you know. But, that's my concern". (P1, stylist)<br><br>"For pills, I am not sure how safe they will be in the salon environment, especially considering temperature requirements, if those would be ideal in a salon setting. [...] But with select products like condoms, I'd have no problem providing them from my salon." (P1, stylist) |
| | Salon not hygienic enough | "I don't know because it's sealed so if I could say the salons are not clean for things that go into our mouths but I remember that they are sealed right? [...] in that way it cannot come into contact with products from the salon but if we ensure that it is safe then there is no problem." (P7, stylist) |
| | Sometimes men in salon | "[My clients] are males and females […] what I've realised is that [talking about family planning at the salon that serves men] I don't know if it could be something that's not offensive or not" (P8, male stylist)<br><br>"Haaii, I can say being a Mosotho, some can agree and be happy to be given such services by men because I've realised that some women say men need to know about this stuff. Therefore I think some would be glad while others would not and would wonder what kind of information I'm talking about now" (P8, male stylist) |
| **Legal concerns** | Concerns about legal implications | "Heii! Is [the birth control pill] not the one that's said to be illegal that can get me in trouble?" (P8, stylist) |
| **Privacy concerns** | Too many clients present | "Sometimes there's a random person sitting in the salon who isn't even there to do her hair. I don't know, but I think of a cubicle in the same salon space which is enough for a private conversation." (P1, stylist) |
| **Negative impact on business** | Concern about upsetting clients by talking about SRH/HIV services | "As I was filling out the surveys, I wondered whether this is really what I would want to do. Because some people may not be comfortable, and may feel like |

| | | |
|---|---|---|
| | | when they come to my salon, I assume things about them and their behaviors.[...] Some may wonder, 'who is she to tell me about my sexual life?' " (P12, stylist)<br><br>"So I think a disadvantage to that could be, let's say I've talked to someone who is not comfortable, it might happen that she never comes back because she may say 'at your salon you keep talking about this and that'. In that way, it could be a disadvantage to people who are not open to talking about these kinds of services. " (P7, stylist) |
| **Concerns about implementation** | Linkage to care | "And also, I'd still love that the Ministry of Health should be involved so that if somebody's results are positive, then they'd know where to get help…" (P1, stylist) |
| | Accreditation | "I think what can help can be training for a specific time then after the training there should be certificates that I can display at the salon so that when they talk they should see that they are talking to someone who is trained to offer them those services." (P7, stylist)<br><br>"Some clients will ask ,'where did you learn this? Are you in the medical field like a nurse?' Some people will listen to nurses and doctors, not to somebody doing hair." (P3, stylist) |
| | Training | "I think the clinics can support me by providing or training me like in counselling. I think it's not everyone who can do it. You have to pass the process. Education on how to handle things like an abused person or one who needs PrEP and also how to handle a person who is infected with STIs." (P3, stylist) |
| | **Facilitators - Stylists** | |
| **Existing infrastructure** | Already provide some SRH services | "What I do at the salon, I always have some pads, not selling, not even giving them the whole package. I'm just giving someone, if a client is sitting down and then says 'Oh, I'm on my period and I didn't have anything', I still have them in the salon. [...]<br>I haven't even considered selling them. I think by giving them that at the time of emergency yes. Somebody didn't plan it. It just happened. So I'm here to rescue with a pad." (P3, stylist) |
| **Positive impact on business** | Existing clients may visit more frequently | "My clients will come back to the salon regularly. And also if it's at a small cost or for free, then I'll get more clients, and the services will be more affordable to more people." (P1, stylist) |

| | | |
|---|---|---|
| | May attract younger clients | "I think there will be more clients like youth. The youth are comfortable where they feel that they are not judged, and where they hear a person talking freely." (P3, stylist) |
| | May attract new clients | "I feel it's going to bring me more clients. One would tell the other that when you go to that salon you will also get this and this." (P7, stylist)<br><br>"To my salon, I think [the hair salon project] will benefit because more clients will be coming." (P3, stylist) |
| **Client comfort** | Less stigmatizing than health facility | "We tell ourselves that at the clinic we just talk and go back home, but when we are in the queues waiting for services, they specify which queue is for which service and one gets scared to queue on the right line because they believe everyone will see what they are at the clinic for." (P3, stylist) |
| | Good stylist/client relationship | "Yes, there are some people [who can come to me for SRH services] Some don't even know me, it's their first time at the salon and they become comfortable and start telling you what happened. Some might need us to chat for some time before they become comfortable to tell you." (P7, stylist) |
| | Non-judgemental space | "A lot of people don't like going to health centres because we come across [healthcare workers' negative] attitudes. Already, [...] we talk about [SRH services] at the salon so I felt like it was an opportunity to help them talk where it's closer." (P6, stylist)<br><br>"The main reason is that at health centres people are scared. It's crowded and we are not comfortable talking to some doctors. So [at the salon] we are friends. [...] Clients who had filled in the survey were already asking when and how they [would] get the services. I told them we hadn't been told yet. I think it's going to be easy for it to happen." (P6, stylist) |
| | Predominantly female space | "the women get more comfortable. Like, they relax when it's 'woman to woman' and when it's a salon space, I guess. They don't feel like it's formal. They have to. You know, sometimes when things are formal, you even feel the need to lie. But when it's just chilled, you become yourself." (P1, stylist) |
| **Client convenience** | Salon has flexible opening hours | "I also think the time at the salon is convenient for everyone. At the health centres, we go on specific times and dates e.g from Monday to Friday maybe 8 am to 4 pm. But at the salon, one goes every time." |

| | | |
|---|---|---|
| | | (P7, stylist) |
| | Salon nearer than facility | "Another thing is I wouldn't wake up early to go to the health centre to test. I think we will have brought it closer to people." (P7, stylist) |
| | Two-in-one service (health and hair) | "... it's not every time that we go to the clinics. I cannot wake up early and not go to work or elsewhere and say I'm going to the clinic to get the injection or any service that we are talking about. I cannot go for those services [specifically] but I would rather go whenever I have other plans that take me to the clinic, maybe if I'm sick or if I have an appointment or checkup. But at the salon one goes almost every month or after 2/3 weeks." (P7, stylist) |

| **Barriers - Clients** | | |
|---|---|---|
| **Themes** | **Codes** | **Illustrative quotes** |
| **Privacy concerns** | Stylist may share personal information | "I feel like things such as PrEP and HIV are very sensitive. You need someone who is very discrete, and someone with whom you will never have a conflict. I would much rather prefer to get it from the clinic. [The clinic] is better than the salon! I know them but I need confidentiality. I fear that if I have a conflict with my stylist, she would tell people about my family affairs." (P11, client) |
| | Salon offers not enough privacy | "And then [the salon has] couches where their next client is expected to sit while waiting for their hairstylist to be done. People coming around, a lot of people doing hair washes going on. They share, like, the room, small room, not so, like, private. So almost everyone can hear what you're saying, what you're talking [about] with your hairstylist." (P14, client)<br><br>"[The other clients might] think I am HIV positive because I asked for an HIV self-test and didn't go to the clinic. Afterwards, they are going to ask me how it had gone. At the clinic, they don't ask you." (P4, client) |
| | Too many other clients present | "If she has clients and she is doing their hair it would be a challenge for me to, like, if she is busy doing hair it will be a challenge for me to talk to her about it and ask for something because at some point it's kind of, like, confidential you know, people are judging." (P10, client) |

| | | |
|---|---|---|
| | Close-knit community | “ [Condoms] should just be where anyone can just take them and pass. Because when you ask for them the stylist might ask many questions [...]<br>Because she knows me as a young girl who attends church with her, she will be shocked at me.” (P2, client) |
| **Implementation concerns** | Stylist needs SRH training | “I know she's somebody that can be, can be able to do it, but just have like, she should also be trained enough to know about them so that she can be able to explain for somebody who doesn't even have little knowledge out there.” (P14, client) |
| | Stylist’ ability to explain side effects | “At the clinic, I think they are going to tell you about the importance of the pill whilst at the salon they may give it to you without explaining its importance and disadvantages. I think I can go to the clinic because they will give me more information about the pill.” (P4, client) |
| | Concerns about product safety | “The challenge would be that I wouldn't know if [the pill or condom at the salon] is safe, is it 100% safe, or is it really [authentic], or not.” (P5, client) |
| | Salon not hygienic enough | “I am very particular with tidiness and cleanliness. So if you're going to start providing health services, I want the space to be clean. So with hair and nails and other women in one room, when are we gonna get the time to talk?” (P9 client) |
| **Client discomfort** | Poor stylist/client relationship | “I know them but I need confidentiality. I fear that if I have a conflict with my stylist, she would tell people about my family affairs.” (P11, client) |
| | **Facilitators - Clients** | |
| **Client comfort** | Non-judgemental space | “Well it's a salon so like I said at the salon I don't think there will be a lot of people who will be judgemental because I think the person who will have placed it there will have a reason why they placed [the pill] there so she will be expecting people to take them.” (P5, client)<br><br>“A top challenge for me is that sometimes I don't want to go to the hospital. Some nurses ask questions that are too judgemental (laughs), yes we are young, and we are already sexually active, but the last thing we need is to be judged.” (P13, client) |
| | Good stylist/client relationship | “It is important to get a hairstylist that you trust... So there needs to be trust between you and your stylist.” (P13, client) |

| | | |
|---|---|---|
| | | “I think I would be comfortable [at the salon]. Because she is not going to talk to me like someone who is uptight, for example talking to a nurse. She is probably going to start following from a conversation we are already having. So it would be a lot more engaging because we will be talking about many other things.” (P13, client) |
| | Predominantly female space | “In the salon, we are women. [Condomless sex] is something that can happen to anyone, and so they would all understand. It’s not a bad thing.” (P11, client) |
| **Client convenience** | Tow-in-one service (health and hair) | “Since women already go to salons almost every month, it would be easier for them to get contraceptives from there.” (P13, client) |
| | Salon’s flexible opening hours | “It's somewhere you can just get ready and go get it without having to worry about having to find them closed. It's somewhere that you can just call, 'I need this. I need this', anytime you want to.” (P14, client) |
| **Comparative advantage over health facilities** | Avoiding queues at health facility | “It saves time because when I get to the clinic I am going to find people there whilst at the salon there might not be a lot of people.” (P4, client)<br><br>“I will be motivated [to get PrEP from my stylist] because it will be closer. I won't need to travel or take a long time to get it or wait in long queues at clinics or hospitals.” (P5, client) |
| | Salon is nearer than health facility | “It could benefit them because walking from my village to the health center is a long distance, so if one goes to a hair salon close by, it wouldn't take long like walking to the health center.” (P2, client)<br><br>“The advantage for getting PrEP [from the salon], I think it would be [that] usually our hair salons, they are close by, [...] you don't have to worry about going to like, pharmacists to get them or anywhere, any other places where you have to like use transport to get to reach to that place, lazy to just bath and go there.” (P14, client) |
| | Less stigmatizing than pharmacy or health facility | “Okay, I'm not trying, like, to generalize, but in [government health facilities] I've heard, I've seen people being judged based on their ages. Normally, for [...] youth who are not yet comfortable enough to expose that they are having ARVs. And I was [...] in an incident where I saw, like, this lady trying to hide those pills, and the comment that they got from the nurses was not, was not very, very, very good, because they |

| | | |
|---|---|---|
| | | were like, ‘okay, if you are young enough to do this (sex), how come you are just afraid to take the pills, yet they are going to protect you?’ [...]<br>So that's why I'm saying to avoid getting all those, I would really recommend going to hair salons.” (P14, client) |
| **Cost considerations** | Salon has credit payment system | “If the prices are the same, we might as well go to the shops. Unless the hair stylist is open with me getting them on credit. Sometimes when I don’t have enough money to do my hair, I write my name down and do it on credit, then I pay later when the month ends.” (P11, client) |

Table S6. Reasons for stylists' discomfort in offering sexual and reproductive health services.

| | | **Privacy issues** | **Feeling unconfident in their stylists' abilities to provide the services.** | **Other** | **Total participants*** |
|---|---|---|---|---|---|
| **HIV** | HIV counselling | 5 (41.67%) | 5 (41.67%) | 3 (25%) | 12 |
| | Oral HIV self-testing | 3 (60%) | 3 (60%) | 0 (0%) | 5 |
| | HIV pre-exposure prophylaxis | 3 (42.86%) | 3 (42.86%) | 1 (14.29%) | 7 |
| | HIV post-exposure prophylaxis | 2 (33.33%) | 3 (50%) | 1 (16.67%) | 6 |
| **Family planning and sexually transmitted infections** | Family planning counselling | 6 (75%) | 2 (25%) | 2 (25%) | 8 |
| | Sexually transmitted infection counselling | 8 (66.67%) | 4 (33.33%) | 2 (16.67%) | 12 |
| | External/male condoms | 5 (83.33%) | 1 (16.67%) | 0 (0%) | 6 |
| | Internal condoms/female condoms | 5 (83.33%) | 1 (16.67%) | 0 (0%) | 6 |
| | Contraceptive pill | 4 (57.14%) | 2 (28.57%) | 1 (14.29%) | 7 |
| | Emergency contraceptive pill | 4 (66.67%) | 1 (16.67%) | 1 (16.67%) | 6 |
| **Gender-based violence** | Gender-based violence information | 4 (80%) | 0 (0%) | 1 (20%) | 5 |
| **Menstrual health** | Menstrual counselling | 3 (60%) | 2 (40%) | 0 (0%) | 5 |
| | Menstrual products | 2 (66.67%) | 1 (33.33%) | 0 (0%) | 3 |

**Note: Participants could select more than one answer.**

*Who (strongly) disagreed with being comfortable providing the respective service.

Table S7. Reasons for clients' discomfort in receiving sexual and reproductive health services.

| | | Privacy issues | Feeling unconfident in their abilities to provide the services | Religious beliefs | Not interested | Prefer professional care | Total participants* |
|---|---|---|---|---|---|---|---|
| **HIV** | HIV counselling | 24 (66.67%) | 7 (19.44%) | 0 (0%) | 0 (0%) | 15 (41.67%) | 36 |
| | Oral HIV self-testing | 4 (28.57%) | 3 (21.43%) | 1 (7.14%) | 0 (0%) | 10 (71.43%) | 14 |
| | HIV pre-exposure profilaxis | 6 (30%) | 7 (35%) | 0 (0%) | 1 (5%) | 14 (70%) | 20 |
| | HIV post-exposure profilaxis | 6 (23.08%) | 8 (30.77%) | 0 (0%) | 0 (0%) | 16 (61.54%) | 26 |
| **Family planning and sexually transmitted infections** | Family planning counselling | 12 (50%) | 5 (20.83%) | 1 (4.17%) | 0 (0%) | 12 (50%) | 24 |
| | Sexually transmitted infection counselling | 18 (52.94%) | 10 (29.41%) | 1 (2.94%) | 1 (2.94%) | 15 (44.12%) | 34 |
| | External/male condoms | 7 (35%) | 3 (15%) | 2 (10%) | 3 (15%) | 7 (35%) | 20 |
| | Internal condoms/female condoms | 7 (33.33%) | 6 (28.57%) | 1 (4.76%) | 1 (4.76%) | 8 (38.1%) | 21 |
| | Contraceptive pill | 11 (40.74%) | 4 (14.81%) | 1 (3.7%) | 2 (7.41%) | 12 (44.44%) | 27 |
| | Emergency contraceptive pill | 8 (40%) | 7 (35%) | 1 (5%) | 1 (5%) | 7 (35%) | 20 |
| **Gender-based violence** | Gender-based violence information | 5 (33.33%) | 7 (46.67%) | 0 (0%) | 1 (6.67%) | 4 (26.67%) | 15 |
| **Menstrual health** | Menstrual counselling | 8 (42.11%) | 6 (31.58%) | 0 (0%) | 0 (0%) | 8 (42.11%) | 19 |
| | Menstrual products | 3 (33.33%) | 1 (11.11%) | 1 (11.11%) | 2 (22.22%) | 3 (33.33%) | 9 |

**Notes:**

- Participants could select more than one answer.

- Respondents had the option to select 'other,' but after review, all responses were reclassified into existing categories.

*Who (strongly) disagreed with being comfortable receiving the respective service.

Table S8. Impact stylists report believing offering SRH services could have on their business.

| | **n = 157** |
|---|---|
| Offering SRH services would have a positive impact on the business (%) | 142 (90.4) |
| Reasons why stylists believe it could positively impact their business. | |
| Attract new clients* (%) | 102 (71.8) |
| Usual clients may come more frequently* (%) | 53 (37.3) |
| Other reason* (%) | 9 ( 6.3) |
| Offering SRH services would have a negative impact on the business (%) | 17 (10.8) |
| Reasons why stylists believe it could negatively impact their business. | |
| I may lose working hours* (%) | 5 (29.4) |
| Clients may not come to my hair salon because they dislike the idea* (%) | 12 (70.6) |
| Other reason* (%) | 1 ( 5.9) |

**Note:**

* Participants could select more than one answer.

Table S9. Accessibility of hair salons in terms of both cost and time.

| n = 308 | | Hair salon | Nearby health facility |
|---|---|---|---|
| Transport time (%) | Less than 15 min | 103 (33.4) | 78 (25.3) |
| | Between 15 and 30 min | 150 (48.7) | 156 (50.6) |
| | Between 30 and 60 min | 40 (13.0) | 57 (18.5) |
| | More than an hour | 15 ( 4.9) | 17 ( 5.5) |
| Transport cost (%) | M0-M10 | 87 (28.2) | 68 (22.1) |
| | M10-M20 | 123 (39.9) | 140 (45.5) |
| | M20-M30 | 68 (22.1) | 76 (24.7) |
| | More than M30 | 30 ( 9.7) | 24 ( 7.8) |
